# Hepatic Mitochondrial Alterations in GLP-1 Poor Responders Undergoing Roux-en-Y Gastric Bypass

**DOI:** 10.64898/2026.08.04.26359613

**Authors:** Akshay Pratap, Breanna Juda, Daniel Bessesen, Adam Gilden, Lindsey Westbrook, John Janetsko, Kenneth Meza Monge, Anza Dareshouri, Juan Pablo Idrovo, Kevin Rothchild, Bryan C Bergman, Nalu Navarro-Alvarez

## Abstract

**Background:** The expanding use of glucagon-like peptide-1 receptor agonists (GLP-1 RAs) has created a growing population of patients undergoing metabolic surgery after inadequate pharmacologic weight-loss response. Whether prior poor response is associated with distinct hepatic metabolic abnormalities or altered postoperative recovery is still unclear.

**Materials and Methods:** In this prospective observational study, 20 patients undergoing Roux-en-Y gastric bypass (RYGB) were enrolled, including 10 patients with GLP-1 receptor agonist–refractory obesity (GRO), operationally defined as less than 5% total body weight loss after at least six months of adequate GLP-1 RA therapy, and 10 GLP-1 RA-naïve patients (GNO). Five lean individuals served as hepatic reference controls. Paired liver biopsies obtained during RYGB and approximately six months postoperatively were evaluated using histology, BODIPY fluorescence, transmission electron microscopy, and Western blot analysis of AMPK–ACC signaling and mitochondrial oxidative phosphorylation proteins.

**Results:** Baseline anthropometric and metabolic characteristics were broadly comparable between surgical cohorts. Nevertheless, GRO exhibited greater hepatic lipid accumulation, higher histological disease activity, more pronounced mitochondrial ultrastructural abnormalities, and reduced AMPK–ACC phosphorylation. Both cohorts achieved substantial six-month postoperative weight loss, although mean percentage total weight loss was lower in GRO than GNO (25.21% versus 29.51%). Paired tissue analyses demonstrated postoperative improvements in hepatic histology, lipid accumulation, and mitochondrial cristae integrity in both cohorts, with smaller improvements in GRO. Postoperative AMPK and ACC phosphorylation ratios remained lower in GRO, while mitochondrial protein analyses demonstrated complex-specific remodeling, including lower SDHB, UQCRC2, COX IV, and ATP5A abundance despite comparable NDUFS8 abundance at six months.

**Conclusions:** Prior poor GLP-1 RA weight-loss response was associated with distinct hepatic histological, energy-sensing, and mitochondrial characteristics among patients undergoing RYGB. Although substantial surgical weight loss occurred in both cohorts, differences in hepatic tissue recovery and molecular profiles persisted at six months. These exploratory findings warrant validation in larger cohorts incorporating pharmacologic responders and direct mitochondrial functional assessments.

**Highlights:**

- Poor GLP-1 receptor agonist response was associated with distinct hepatic metabolic abnormalities.
- Paired human liver biopsies enabled longitudinal assessment of hepatic remodeling after RYGB.
- GRO patients exhibited greater hepatic steatosis, mitochondrial disruption, and altered AMPK–ACC signaling.
- Both cohorts achieved substantial weight loss, but hepatic histological and mitochondrial recovery was less pronounced in GRO.
- Complex-specific mitochondrial OXPHOS remodeling persisted six months after RYGB, despite postoperative improvement.

## Introduction

Glucagon-like peptide-1 receptor agonists (GLP-1 RAs) have substantially expanded pharmacologic treatment options for obesity and associated metabolic disorders.^1, 2^ Large clinical trials have demonstrated clinically meaningful weight reduction and improvements in cardiometabolic outcomes with GLP-1-based therapies.^3, 4^ Nevertheless, treatment response remains heterogeneous, and a subset of patients experiences limited weight loss despite prolonged pharmacologic treatment.^4, 5^ For the purposes of this study, patients with inadequate weight-loss response to GLP-1 RA therapy are designated as having GLP-1 receptor agonist– refractory obesity (GRO). This operational classification refers to failure to achieve at least 5% total body weight loss despite a minimum of six months of adequate treatment and does not imply established biological resistance to GLP-1 receptor signaling. As GLP-1 RA use increases, metabolic and bariatric surgeons are increasingly likely to evaluate patients with a history of inadequate response to these medications. Whether poor response to pharmacologic treatment is associated with subsequent weight-loss outcomes after bariatric surgery remains incompletely understood.

Roux-en-Y gastric bypass (RYGB) produces substantial and sustained weight loss through coordinated changes in gastrointestinal physiology, nutrient handling, appetite regulation, and metabolic signaling.^6, 7^ The operation also improves metabolic dysfunction-associated steatotic liver disease (MASLD), including hepatic steatosis and, in some patients, fibrosis.^8^ However, postoperative weight loss and metabolic recovery vary among individuals. Whether previous response to GLP-1 RA therapy identifies patients with differences in surgical outcomes or underlying metabolic characteristics is an important clinical question. Identifying such associations could improve understanding of treatment-response heterogeneity and guide future investigations into the biological determinants of surgical outcomes.

The liver provides an opportunity to investigate metabolic differences associated with treatment response. Hepatic energy sensing, mitochondrial biogenesis, and oxidative phosphorylation are central to lipid metabolism and metabolic homeostasis.^9^ AMP-activated protein kinase (AMPK) regulates multiple metabolic pathways, including fatty acid metabolism and mitochondrial homeostasis.^10, 11^ Alterations in these pathways have been implicated in hepatic steatosis and metabolic dysfunction.^12^ Although GLP-1-based therapies may influence hepatic metabolism through direct or indirect mechanisms^13^, the relationship between prior pharmacologic weight-loss response and hepatic molecular characteristics in patients undergoing bariatric surgery has not been adequately characterized.

Importantly, inadequate response to GLP-1 RA therapy may coexist with differences in underlying insulin resistance, metabolic disease severity, or other patient-specific characteristics. Therefore, alterations in hepatic molecular signaling cannot necessarily be attributed to impaired GLP-1 receptor activity or pharmacologic nonresponse itself. Integrating clinical outcomes with hepatic histology, mitochondrial ultrastructure, and molecular profiling may help identify biological characteristics associated with treatment-response heterogeneity.^12^ Liver tissue obtained during bariatric surgery provides a valuable opportunity to investigate these relationships directly in a clinically relevant population.^14, 15^

We therefore conducted a prospective observational study comparing patients with prior poor response to GLP-1 RA therapy and GLP-1 RA-naïve patients undergoing RYGB. The objectives were to (1) compare postoperative weight-loss outcomes, (2) characterize differences in baseline hepatic histology, mitochondrial ultrastructure, and metabolic protein expression, and (3) evaluate longitudinal changes in hepatic characteristics using paired liver biopsies obtained from all surgical participants. We additionally examined exploratory associations between baseline hepatic protein expression and postoperative weight loss. We hypothesized that patients with prior poor pharmacologic response would exhibit differences in postoperative weight loss and hepatic metabolic characteristics compared with GLP-1 RA-naïve patients.

## Materials and Methods

### Study Design and Participants

This prospective observational cohort study was conducted at the University of Colorado Anschutz Medical Center following approval by the Colorado Multiple Institutional Review Board (COMIRB Protocol 25-0802). Written informed consent was obtained from all participants. The study was conducted in accordance with the Declaration of Helsinki and reported following the Strengthening the Reporting of Observational Studies in Epidemiology (STROBE) guidelines. Between January 2025 and July 2026, 46 adults undergoing evaluation for primary Roux-en-Y gastric bypass (RYGB) were screened for eligibility. Twenty patients with obesity were prospectively enrolled into two cohorts: patients with prior poor response to glucagon-like peptide-1 receptor agonist therapy (GRO; n = 10) and GLP-1 receptor agonist-naïve patients (GNO; n = 10). GNO participants were selected to achieve group-level comparability with GRO participants for baseline body mass index (BMI), type 2 diabetes prevalence, and insulin resistance, assessed using the homeostatic model assessment of insulin resistance (HOMA-IR). Individual patient matching was not performed. Five metabolically healthy lean adults undergoing elective abdominal surgery were included as physiological reference controls. Lean specimens were used for cross-sectional hepatic comparisons and were not included in the primary clinical outcome or longitudinal paired analyses. Eligible surgical participants were aged ≥18 years with BMI ≥35 kg/m² and at least one obesity-related comorbidity or BMI ≥40 kg/m². Exclusion criteria included prior bariatric surgery, type 1 diabetes, chronic corticosteroid therapy, endocrine or genetic disorders affecting weight regulation, and inability to tolerate adequate GLP-1 receptor agonist treatment when applicable.

### GLP-1 Receptor Agonist Exposure and Definition of Poor Response

GLP-1 receptor agonist–refractory obesity (GRO) was operationally defined as less than 5% total body weight loss following at least six months of treatment at the prescribed maintenance dose after completion of standard dose escalation, with adherence documented through clinical records. GRO represents a study-specific clinical classification based on observed pharmacologic weight-loss response rather than an established diagnosis or demonstrated impairment of GLP-1 receptor signaling. The GRO cohort included eight patients previously treated with semaglutide (2.4 mg weekly) and two treated with liraglutide (3.0 mg daily). Treatment duration, maintenance dosing, adherence, and pharmacologic weight-loss response were assessed using electronic health records, prescription histories, and clinician documentation. The GLP-1 RA naïve (GNO) cohort comprised of patients without prior GLP-1 receptor agonist exposure. GLP-1 receptor agonist therapy was withheld for seven days before surgery according to the institutional perioperative medication-management protocol. GRO was used as an operational clinical classification rather than an established biological disease entity. Additional treatment-exposure characteristics are presented in Table 2.

### Surgical Procedure and Longitudinal Liver Biopsy Collection

All 20 surgical participants underwent standardized primary RYGB using a 120-cm biliopancreatic limb and a 100-cm Roux limb. All participants had documented gastroesophageal reflux disease, which contributed to the clinical selection of RYGB. As part of the institutional surgical pathway, staged cholecystectomy was prospectively planned approximately 6–8 months after RYGB. This institution-specific strategy was incorporated into preoperative counseling and informed consent. The research protocol incorporated postoperative liver biopsy collection during the planned procedure. Baseline liver biopsies (T0) were obtained during the index RYGB, and postoperative biopsies (T6) were collected during staged cholecystectomy. All 20 participants completed both biopsy time points, yielding 20 matched biopsy pairs (10 GNO and 10 GRO). Specimens were allocated for histological examination, fluorescence-based lipid quantification, transmission electron microscopy (TEM), and Western blot analysis. Detailed tissue-processing procedures are provided in the Supplementary Methods.

### Hepatic Histology and Mitochondrial Ultrastructural Assessment

Hepatic histological disease activity was evaluated using hematoxylin and eosin staining and the nonalcoholic fatty liver disease activity score (NAS; range, 0–8). Neutral lipid accumulation was assessed using BODIPY 493/503 fluorescence staining, with lipid-positive area expressed as a percentage of evaluable tissue area. Mitochondrial ultrastructure was examined using TEM. Cristae integrity was assessed using a semiquantitative 0–4 scoring system, with higher scores indicating greater structural preservation. Ten non-overlapping TEM fields were analyzed per biopsy, with five mitochondrial profiles scored per field. Microscopic measurements were aggregated at the participant level before statistical analysis. Detailed histological scoring criteria, fluorescence image analysis, and mitochondrial morphometry procedures are described in the Supplementary Methods.

### Western Blot Analysis

Hepatic energy-sensing signaling and mitochondrial oxidative phosphorylation (OXPHOS) protein abundance were assessed by Western blotting. Energy-sensing proteins included phosphorylated and total AMP-activated protein kinase (pAMPK/AMPK) and acetyl-CoA carboxylase (pACC/ACC). Mitochondrial OXPHOS proteins included NDUFS8 (complex I), SDHB (complex II), UQCRC2 (complex III), COX IV (complex IV), and ATP5A (complex V). Protein abundance was quantified by densitometry. Phosphorylated proteins were normalized to their corresponding total proteins, whereas OXPHOS proteins were normalized to the designated loading controls. Five lean reference specimens and 10 specimens from each surgical cohort were included in baseline Western blot analyses. Detailed immunoblotting and densitometric procedures are provided in the Supplementary Methods.

### Clinical Outcomes

Demographic characteristics, anthropometric measurements, metabolic comorbidities, medication exposure, and laboratory parameters were obtained from the institutional bariatric surgery registry and electronic health records. The primary clinical outcome was percentage total body weight loss (%TWL) at six months following RYGB, calculated as:

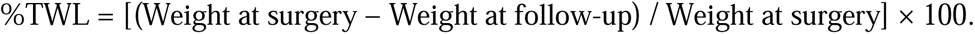

Three-month %TWL and percentage excess BMI loss (%EBMIL) were evaluated as secondary outcomes.

### Statistical Analysis

Continuous variables were summarized as mean ± standard deviation (SD) or median [interquartile range (IQR)], as appropriate, and categorical variables as number (percentage). Baseline clinical characteristics were compared between GRO and GNO using two-sided Welch’s independent-samples t-tests for continuous variables and Fisher’s exact tests for categorical variables, as appropriate. Cross-sectional comparisons of hepatic AMPK–ACC phosphorylation ratios and mitochondrial oxidative phosphorylation (OXPHOS) protein abundance among lean reference controls, GNO, and GRO were performed using ordinary one-way analysis of variance (ANOVA), followed by Sidak-adjusted multiple comparisons for lean versus GNO, lean versus GRO, and GNO versus GRO for AMPK-ACC and OXPHOS outcomes. Baseline histological and ultrastructural outcomes were analyzed using ordinary one-way ANOVA followed by Šídák’s multiple-comparisons test for the three pairwise comparisons (lean versus GNO, lean versus GRO, and GNO versus GRO). Longitudinal hepatic outcomes were evaluated using matched baseline (T0) and postoperative (T6) measurements. Within-participant changes were calculated as Δ = T6 − T0. Between-group differences in participant-level changes in hepatic histological disease activity, intrahepatic lipid accumulation, and mitochondrial cristae integrity were assessed using two-sided unpaired Student’s t-tests assuming equal variances. Between-group differences in changes in AMPK–ACC phosphorylation ratios and OXPHOS protein abundance were assessed using two-sided Welch’s independent-samples t-tests. Individual microscopic fields and mitochondrial profiles were treated as nested observations and aggregated at the participant level before statistical comparisons to avoid pseudoreplication. Within-cohort repeated-measures analyses were performed where appropriate for matched observations. Postoperative percentage total body weight loss (%TWL) and percentage excess body mass index loss (%EBMIL) were compared between cohorts at three and six months using two-sided Welch’s unpaired t-tests in GraphPad Prism, with between-group differences expressed in percentage points (GNO − GRO) and corresponding 95% confidence intervals. Analyses were unadjusted for baseline covariates. Individual participant values are displayed in histological and molecular figures, with group summaries presented as mean ± standard error of the mean (SEM), unless otherwise specified. All tests were two-sided, with nominal statistical significance defined as P < 0.05. Given the small sample size and multiple histological and molecular endpoints, analyses were considered exploratory, and no global adjustment for multiple comparisons was applied. Additional analytical details are provided in the Supplementary Methods.

## Results

### Patient Characteristics and Baseline Metabolic Profile

Between January 2025 and July 2026, 46 patients undergoing evaluation for Roux-en-Y gastric bypass (RYGB) were screened for study participation (Figure 1). Of these, 26 were not enrolled: 10 declined participation and 16 were excluded because they were undergoing revisional bariatric surgery. The remaining 20 patients were prospectively enrolled, comprising 10 patients with prior poor response to GLP-1 receptor agonist therapy (GRO) and 10 GLP-1 receptor agonist-naïve patients (GNO). Five metabolically healthy lean individuals undergoing elective abdominal surgery were included separately as physiological reference controls. All 20 surgical participants completed baseline (T0) and postoperative (T6) liver biopsy collection, yielding 20 matched biopsy pairs for longitudinal analysis.

**Figure 1.**
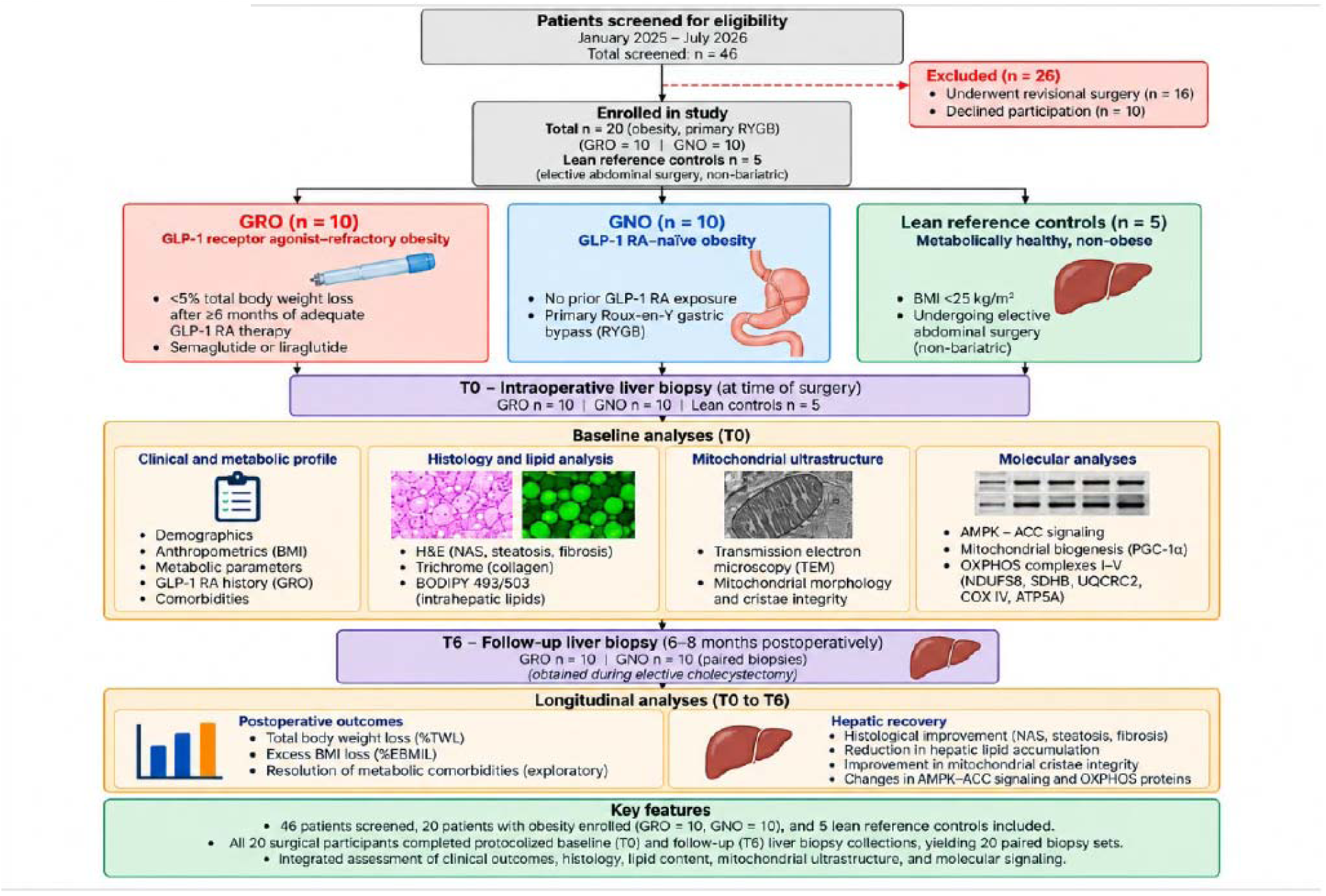
Study Design, Patient Enrollment, and Longitudinal Hepatic Assessments Following Roux-en-Y Gastric Bypass. Schematic overview of the prospective observational study conducted at the University of Colorado Anschutz Medical Center between January 2025 and July 2026. Of 46 patients screened for eligibility, 20 patients with obesity were enrolled and underwent primary Roux-en-Y gastric bypass (RYGB), including 10 patients with GLP-1 receptor agonist–refractory obesity (GRO), operationally defined as less than 5% total body weight loss following at least six months of adequate GLP-1 receptor agonist (GLP-1 RA) therapy, and 10 GLP-1 RA-naïv patients (GNO). Five metabolically healthy lean individuals undergoing elective abdominal surgery served as physiological reference controls. Baseline (T0) liver biopsies were obtained intraoperatively from all 25 participants. Hepatic assessments included histological disease activity using the nonalcoholic fatty liver disease activity score (NAS), intrahepatic neutral lipid accumulation using BODIPY 493/503 fluorescence staining, mitochondrial ultrastructure and cristae integrity using transmission electron microscopy (TEM), and immunoblot analysis of AMPK–ACC signaling and mitochondrial oxidative phosphorylation (OXPHOS) proteins. Postoperative (T6) liver biopsies were obtained approximately 6–8 months after RYGB during protocolized staged cholecystectomy in all 20 surgical participants, yielding 20 matched biopsy pairs (GRO, n = 10; GNO, n = 10). Longitudinal analyses evaluated postoperative percentage total body weight loss (%TWL), percentage excess body mass index loss (%EBMIL), hepatic histological and lipid changes, mitochondrial cristae integrity, and AMPK–ACC and OXPHOS protein remodeling. Lean controls were included exclusively in baseline hepatic reference comparisons and did not undergo longitudinal assessment. Abbreviations: ACC, acetyl-CoA carboxylase; AMPK, AMP-activated protei kinase; BMI, body mass index; GNO, GLP-1 receptor agonist–naïve obesity; GRO, GLP-1 receptor agonist– refractory obesity; NAS, nonalcoholic fatty liver disease activity score; OXPHOS, oxidative phosphorylation; RYGB, Roux-en-Y gastric bypass; T0, baseline; T6, postoperative follow-up; TEM, transmission electro microscopy; %EBMIL, percentage excess body mass index loss; %TWL, percentage total body weight loss.

### Baseline demographic and anthropometric characteristics

The GRO and GNO cohorts were comparable in age (47.2 ± 8.3 versus 45.6 ± 7.8 years; P = 0.662) and preoperative body mass index (BMI; 53.8 ± 2.9 versus 54.2 ± 3.1 kg/m²; P = 0.769). Female participants accounted for 50% of the GRO cohort and 40% of the GNO cohort (P = 1.000). The proportion of Hispanic or Latino participants was identical between groups (30% each; P = 1.000). The distribution of reported racial categories was also similar, with White participants accounting for 50% of GRO and 60% of GNO (P = 1.000), and Black or African American participants accounting for 20% of each cohort (P = 1.000). The lean reference group had a substantially lower BMI (23.1 ± 2.1 kg/m²) and a mean age of 22.4 ± 3.1 years (Table 1).

**Table 1.**
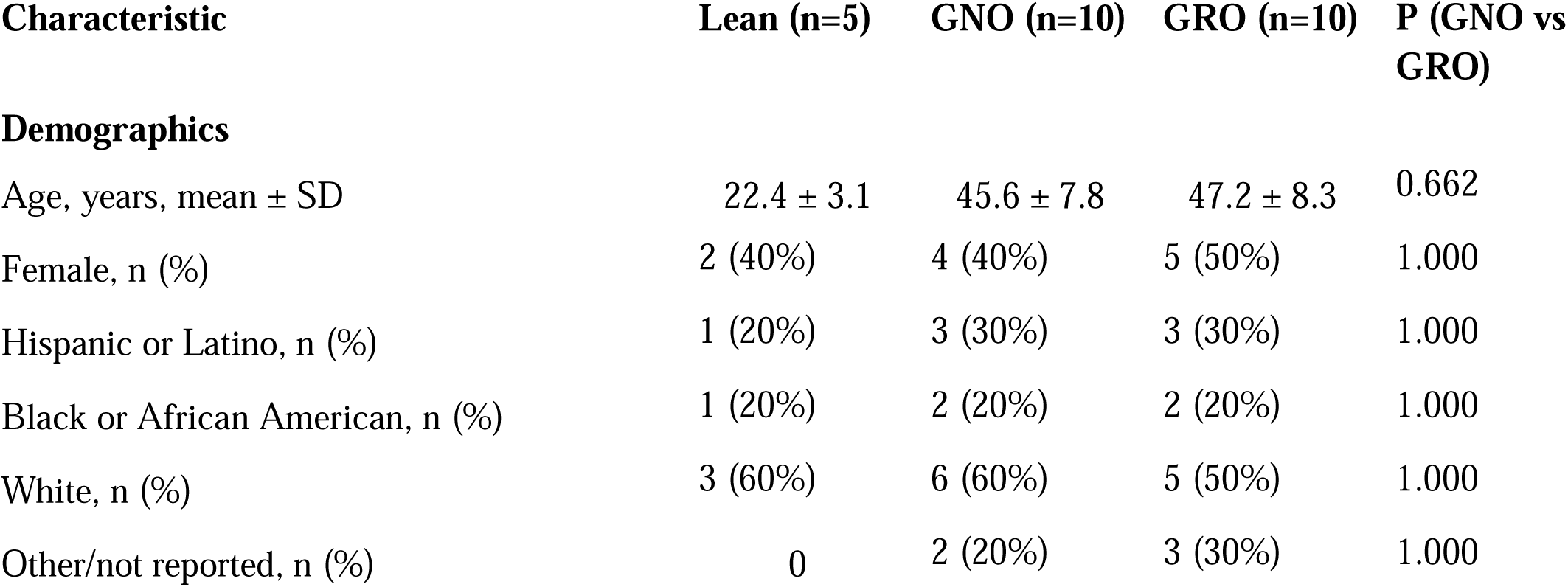
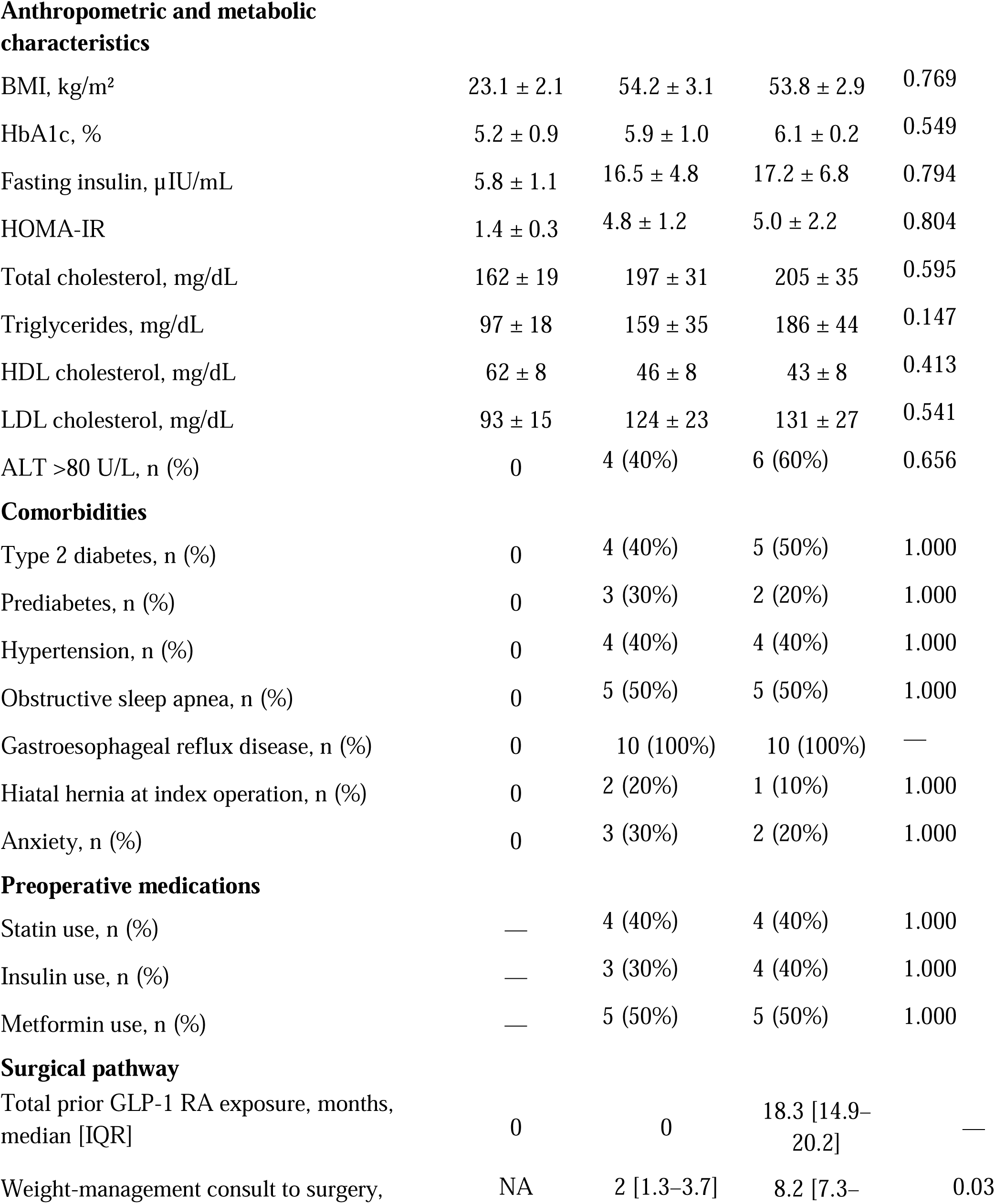
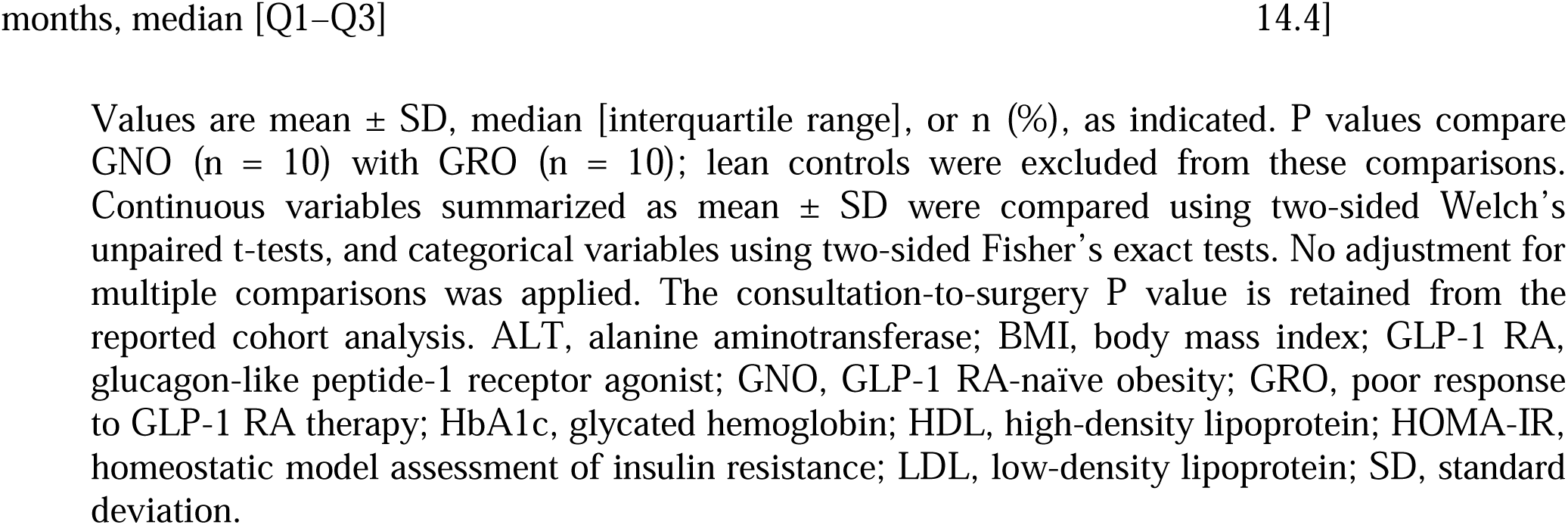
Baseline Demographic, Clinical, and Metabolic Characteristics of Patients With Prior Poor Response to GLP-1 Receptor Agonist Therapy, GLP-1 RA-Naïve Patients, and Lean Reference Controls.

| Characteristic | Lean (n=5) | GNO (n=10) | GRO (n=10) | P (GNO vs GRO) |
| --- | --- | --- | --- | --- |
| **Demographics** |  |  |  |  |
| Age, years, mean ± SD | 22.4 ± 3.1 | 45.6 ± 7.8 | 47.2 ± 8.3 | 0.662 |
| Female, n (%) | 2 (40%) | 4 (40%) | 5 (50%) | 1.000 |
| Hispanic or Latino, n (%) | 1 (20%) | 3 (30%) | 3 (30%) | 1.000 |
| Black or African American, n (%) | 1 (20%) | 2 (20%) | 2 (20%) | 1.000 |
| White, n (%) | 3 (60%) | 6 (60%) | 5 (50%) | 1.000 |
| Other/not reported, n (%) | 0 | 2 (20%) | 3 (30%) | 1.000 |
| **Anthropometric and metabolic characteristics** |  |  |  |  |
| BMI, kg/m<sup>2</sup> | 23.1 ± 2.1 | 54.2 ± 3.1 | 53.8 ± 2.9 | 0.769 |
| HbA1c, % | 5.2 ± 0.9 | 5.9 ± 1.0 | 6.1 ± 0.2 | 0.549 |
| Fasting insulin, µIU/mL | 5.8 ± 1.1 | 16.5 ± 4.8 | 17.2 ± 6.8 | 0.794 |
| HOMA-IR | 1.4 ± 0.3 | 4.8 ± 1.2 | 5.0 ± 2.2 | 0.804 |
| Total cholesterol, mg/dL | 162 ± 19 | 197 ± 31 | 205 ± 35 | 0.595 |
| Triglycerides, mg/dL | 97 ± 18 | 159 ± 35 | 186 ± 44 | 0.147 |
| HDL cholesterol, mg/dL | 62 ± 8 | 46 ± 8 | 43 ± 8 | 0.413 |
| LDL cholesterol, mg/dL | 93 ± 15 | 124 ± 23 | 131 ± 27 | 0.541 |
| ALT >80 U/L, n (%) | 0 | 4 (40%) | 6 (60%) | 0.656 |
| **Comorbidities** |  |  |  |  |
| Type 2 diabetes, n (%) | 0 | 4 (40%) | 5 (50%) | 1.000 |
| Prediabetes, n (%) | 0 | 3 (30%) | 2 (20%) | 1.000 |
| Hypertension, n (%) | 0 | 4 (40%) | 4 (40%) | 1.000 |
| Obstructive sleep apnea, n (%) | 0 | 5 (50%) | 5 (50%) | 1.000 |
| Gastroesophageal reflux disease, n (%) | 0 | 10 (100%) | 10 (100%) | — |
| Hiatal hernia at index operation, n (%) | 0 | 2 (20%) | 1 (10%) | 1.000 |
| Anxiety, n (%) | 0 | 3 (30%) | 2 (20%) | 1.000 |
| **Preoperative medications** |  |  |  |  |
| Statin use, n (%) | — | 4 (40%) | 4 (40%) | 1.000 |
| Insulin use, n (%) | — | 3 (30%) | 4 (40%) | 1.000 |
| Metformin use, n (%) | — | 5 (50%) | 5 (50%) | 1.000 |
| **Surgical pathway** |  |  |  |  |
| Total prior GLP-1 RA exposure, months, median [IQR] | 0 | 0 | 18.3 [14.9–20.2] | — |
| Weight-management consult to surgery, | NA | 2 [1.3–3.7] | 8.2 [7.3– | 0.03 |
| months, median [Q1–Q3] |  |  | 14.4] |  |

Values are mean ± SD, median [interquartile range], or n (%), as indicated. P values compare GNO (n = 10) with GRO (n = 10); lean controls were excluded from these comparisons. Continuous variables summarized as mean ± SD were compared using two-sided Welch's unpaired t-tests, and categorical variables using two-sided Fisher's exact tests. No adjustment for multiple comparisons was applied. The consultation-to-surgery P value is retained from the reported cohort analysis. ALT, alanine aminotransferase; BMI, body mass index; GLP-1 RA, glucagon-like peptide-1 receptor agonist; GNO, GLP-1 RA-naïve obesity; GRO, poor response to GLP-1 RA therapy; HbA1c, glycated hemoglobin; HDL, high-density lipoprotein; HOMA-IR, homeostatic model assessment of insulin resistance; LDL, low-density lipoprotein; SD, standard deviation.

### Baseline metabolic characteristics

Measures of glycemic regulation and insulin resistance were similar between the two surgical cohorts. Mean glycated hemoglobin (HbA1c) was 6.1 ± 0.2% in GRO compared with 5.9 ± 1.0% in GNO (P = 0.549). Fasting insulin concentrations were 17.2 ± 6.8 and 16.5 ± 4.8 µIU/mL, respectively (P = 0.794), while homeostatic model assessment of insulin resistance (HOMA-IR) values were 5.0 ± 2.2 and 4.8 ± 1.2 (P = 0.804). These findings indicated no statistically significant differences in the measured indices of baseline glycemic control or insulin resistance between the surgical cohorts. Lipid profiles were also comparable. Total cholesterol concentrations were 205 ± 35 mg/dL in GRO and 197 ± 31 mg/dL in GNO (P = 0.595), while triglyceride concentrations were 186 ± 44 and 159 ± 35 mg/dL, respectively (P = 0.147). High-density lipoprotein cholesterol was 43 ± 8 mg/dL in GRO compared with 46 ± 8 mg/dL in GNO (P = 0.413), and low-density lipoprotein cholesterol was 131 ± 27 versus 124 ± 23 mg/dL (P = 0.541). Elevated alanine aminotransferase (>80 U/L) was observed in six GRO patients (60%) and four GNO patients (40%; P = 0.656). Although several metabolic measures were numerically less favorable in GRO, none of these between-group comparisons reached nominal statistical significance (Table 1).

### Obesity-related comorbidities and medication exposure

The prevalence of type 2 diabetes mellitus was 50% in GRO and 40% in GNO (P = 1.000), while prediabetes was present in 20% and 30%, respectively (P = 1.000). Hypertension was present in four patients (40%) in each cohort, and obstructive sleep apnea was documented in five patients (50%) per group. All 20 surgical participants had documented gastroesophageal reflux disease, which contributed to the clinical selection of RYGB. Hiatal hernia was identified at the index operation in one GRO patient (10%) and two GNO patients (20%; P = 1.000).

Anxiety was reported in two GRO patients (20%) and three GNO patients (30%; P = 1.000). Preoperative medication exposure was similar between cohorts. Statins were used by four patients (40%) in each group, while insulin therapy was documented in four GRO patients (40%) and three GNO patients (30%). Metformin use was identical between cohorts, with five patients (50%) in each group receiving treatment. No statistically significant differences were identified in the reported frequencies of these comorbidities or medication exposures (Table 1).

### Preoperative GLP-1 receptor agonist treatment characteristics

Among the 10 GRO patients, eight (80%) had received semaglutide and two (20%) had received liraglutide. The reported maintenance doses were 2.4 mg weekly for semaglutide and 3.0 mg daily for liraglutide. Median total treatment duration was 18.32 months (interquartile range [IQR], 14.90–20.20 months). All GRO participants completed dose escalation and maintained the prescribed maintenance dose for at least six months, with treatment adherence documented in the clinical records. The GRO cohort was defined by less than 5% total body weight loss during adequate GLP-1 RA therapy. No participant achieved the 5% weight-loss threshold. All 10 patients discontinued GLP-1 RA treatment before surgery; the institutional protocol specified a seven-day preoperative hold, although individual hold intervals were not verified, and no postoperative GLP-1 RA resumption was reported during the first six months (Table 2).

**Table 2.**
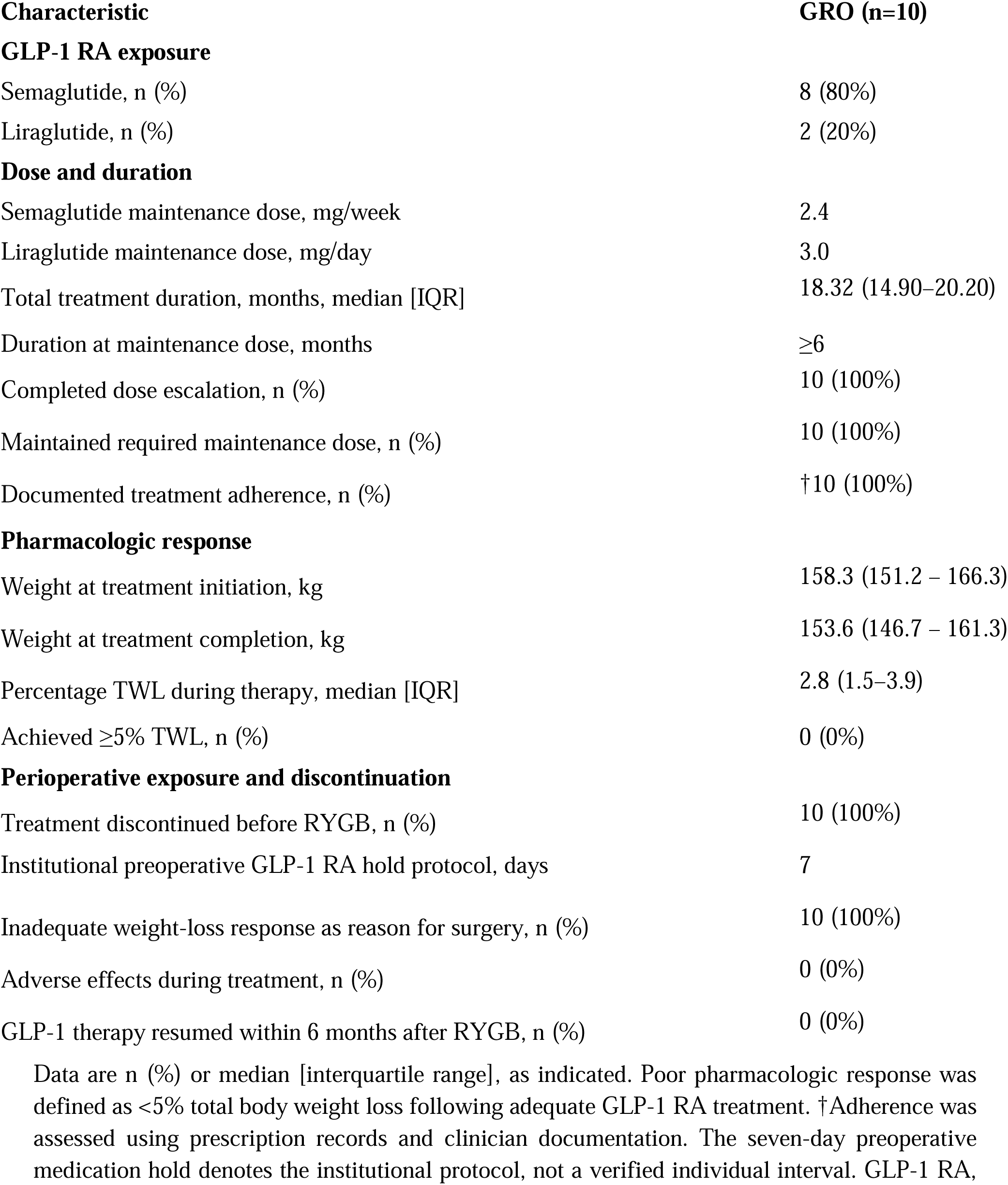
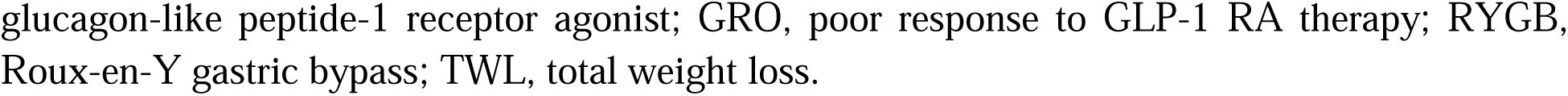
Preoperative GLP-1 Receptor Agonist Treatment Characteristics in Patients with Poor Pharmacologic Response.

| **Characteristic** | **GRO (n=10)** |
| --- | --- |
| **GLP-1 RA exposure** |  |
| Semaglutide, n (%) | 8 (80%) |
| Liraglutide, n (%) | 2 (20%) |
| **Dose and duration** |  |
| Semaglutide maintenance dose, mg/week | 2.4 |
| Liraglutide maintenance dose, mg/day | 3.0 |
| Total treatment duration, months, median [IQR] | 18.32 (14.90–20.20) |
| Duration at maintenance dose, months | ≥6 |
| Completed dose escalation, n (%) | 10 (100%) |
| Maintained required maintenance dose, n (%) | 10 (100%) |
| Documented treatment adherence, n (%) | †10 (100%) |
| **Pharmacologic response** |  |
| Weight at treatment initiation, kg | 158.3 (151.2 – 166.3) |
| Weight at treatment completion, kg | 153.6 (146.7 – 161.3) |
| Percentage TWL during therapy, median [IQR] | 2.8 (1.5–3.9) |
| Achieved ≥5% TWL, n (%) | 0 (0%) |
| **Perioperative exposure and discontinuation** |  |
| Treatment discontinued before RYGB, n (%) | 10 (100%) |
| Institutional preoperative GLP-1 RA hold protocol, days | 7 |
| Inadequate weight-loss response as reason for surgery, n (%) | 10 (100%) |
| Adverse effects during treatment, n (%) | 0 (0%) |
| GLP-1 therapy resumed within 6 months after RYGB, n (%) | 0 (0%) |

Data are n (%) or median [interquartile range], as indicated. Poor pharmacologic response was defined as <5% total body weight loss following adequate GLP-1 RA treatment. †Adherence was assessed using prescription records and clinician documentation. The seven-day preoperative medication hold denotes the institutional protocol, not a verified individual interval. GLP-1 RA,
glucagon-like peptide-1 receptor agonist; GRO, poor response to GLP-1 RA therapy; RYGB, Roux-en-Y gastric bypass; TWL, total weight loss.

### Preoperative surgical pathway

The median interval from initial weight-management consultation to surgery was longer in GRO than GNO patients (8.2 [IQR, 7.3–14.4] versus 2.0 [IQR, 1.3–3.7] months; reported P = 0.03). This difference occurred in the context of prolonged prior pharmacologic treatment among GRO participants, although the observational study design does not establish whether medication exposure directly contributed to the longer surgical pathway. The GRO and GNO cohorts demonstrated no statistically significant differences in the measured baseline anthropometric, metabolic, or clinical characteristics, although the small sample size limits conclusions regarding between-group comparability.

### Baseline Hepatic Histology and Mitochondrial Ultrastructure

Baseline hepatic histological examination revealed distinct differences in liver morphology and lipid accumulation among lean reference controls, GLP-1 receptor agonist-naïve patients (GNO), and patients with prior poor response to GLP-1 receptor agonist therapy (GRO) (Figure 2). Hematoxylin and eosin (H&E) staining demonstrated relatively preserved hepatic architecture in lean controls, whereas GNO specimens exhibited increased hepatocellular vacuolization consistent with steatosis. These abnormalities were more pronounced in GRO, with greater hepatocellular lipid vacuolization and architectural alterations. Quantitative histological assessment demonstrated a higher nonalcoholic fatty liver disease activity score (NAS) in GRO compared with GNO and lean controls (Figure 2). BODIPY 493/503 fluorescence staining further demonstrated differences in intrahepatic neutral lipid accumulation. Minimal lipid-associated fluorescence was observed in lean liver specimens, whereas GNO samples exhibited increased lipid droplet accumulation. GRO specimens demonstrated more extensive and densely distributed lipid-associated fluorescence. Quantitative image analysis confirmed greater BODIPY-positive area in GRO compared with GNO and lean controls (Figure 2). Transmission electron microscopy (TEM) revealed progressive alterations in hepatic mitochondrial ultrastructure across the study cohorts. Mitochondria from lean controls exhibited relatively preserved morphology with organized cristae and intact matrix architecture. GNO specimens demonstrated moderate ultrastructural alterations, including reduced cristae organization in some mitochondria. In contrast, GRO samples exhibited more pronounced mitochondrial abnormalities, characterized by swelling, disrupted cristae, and areas of reduced matrix electron density. Quantitative analysis demonstrated lower mitochondrial cristae integrity scores in GRO compared with GNO and lean controls (Figure 2). Collectively, these findings demonstrate that poor preoperative response to GLP-1 receptor agonist therapy was associated with greater baseline hepatic lipid accumulation, increased histological disease activity, and more pronounced mitochondrial ultrastructural abnormalities at the time of RYGB.

**Figure 2.**
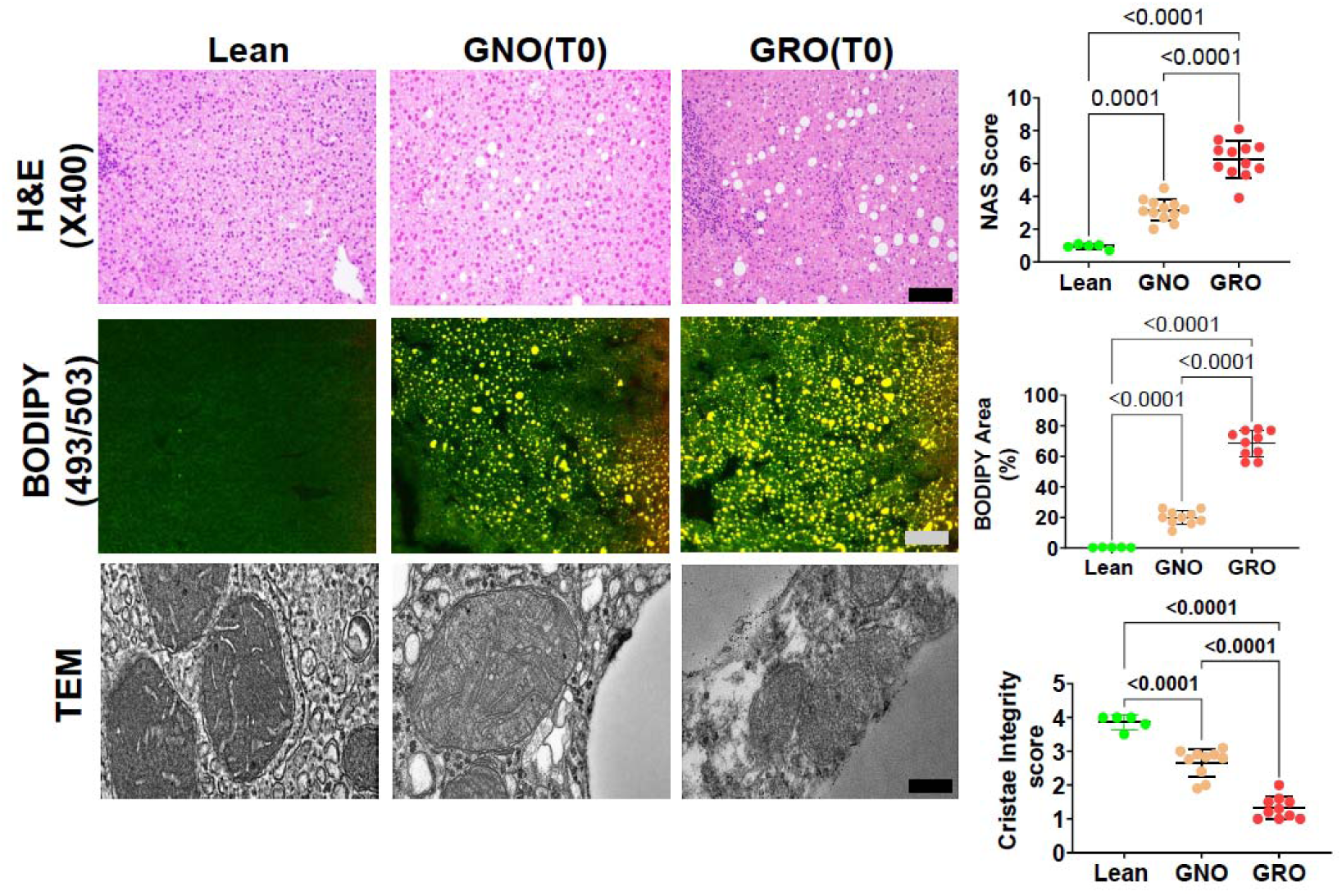
Baseline Hepatic Histological and Mitochondrial Ultrastructural Alterations in Patients with Poo Response to GLP-1 Receptor Agonist Therapy. Representative hematoxylin and eosin (H&E)-stained liver sections (original magnification, ×400), BODIPY 493/503 fluorescence images, and transmission electron micrographs (TEM) from lean physiological reference controls (n = 5), GLP-1 receptor agonist-naïve patients (GNO; n = 10), and patients with prior poor response to GLP-1 receptor agonist therapy (GRO; n = 10) at baseline (T0). Quantitative analyses demonstrate nonalcoholic fatty liver disease activity score (NAS; upper panel), BODIPY-positive area expressed as a percentage of analyzed tissue area (middle panel), and mitochondrial cristae integrity score (lower panel). Individual data points represent independent biological samples. Statistical comparisons were performed using ordinary one-way ANOVA followed by Šídák’s multiple-comparisons test for all three pairwise comparisons. Statistical significance is indicated by **P < 0.01, ***P < 0.001, and ****P < 0.0001. Scale bars: 50 µm (H&E and BODIPY) and 500 nm (TEM).

### Baseline Hepatic AMPK–ACC Signaling and Mitochondrial Oxidative Phosphorylation

To investigate hepatic energy-sensing signaling associated with prior poor response to GLP-1 receptor agonist therapy, we examined AMPK and ACC phosphorylation in baseline liver specimens from lean reference controls, GLP-1 receptor agonist-naïve patients (GNO), and patients with prior poor response to GLP-1 receptor agonist therapy (GRO) (Figure 3). The pAMPK/AMPK ratio was significantly lower in GRO compared with GNO (P < 0.0001) and lean controls (P = 0.0030), whereas no significant difference was observed between GNO and lean controls (P = 0.7773). Similarly, the pACC/ACC ratio was lower in GRO than in GNO (P < 0.0001) and lean controls (P = 0.0055). In contrast, pACC/ACC was higher in GNO than in lean controls (P = 0.0026). These findings indicate that prior poor pharmacologic weight-loss response was associated with reduced hepatic AMPK and ACC phosphorylation at baseline.

**Figure 3.**
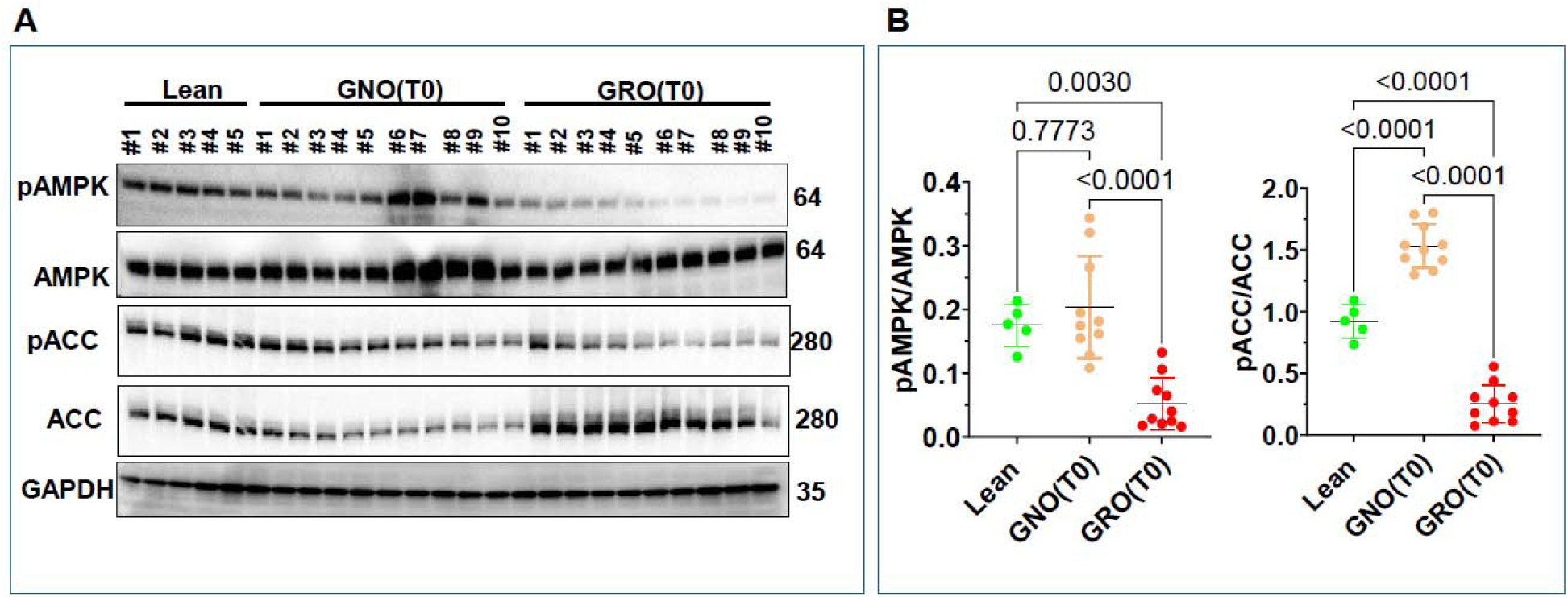
Reduced Hepatic AMPK–ACC Phosphorylation in Patients With Prior Poor Response to GLP-Receptor Agonist Therapy. (A) Representative Western blots showing phosphorylated and total AMP-activated protein kinase (pAMPK and AMPK), phosphorylated and total acetyl-CoA carboxylase (pACC and ACC), an glyceraldehyde-3-phosphate dehydrogenase (GAPDH) in baseline (T0) liver specimens from lean physiological reference controls (n = 5), GLP-1 receptor agonist-naïve patients (GNO; n = 10), and patients with prior poor response to GLP-1 receptor agonist therapy (GRO; n = 10). Molecular weights are indicated in kilodaltons (kDa). (B) Densitometric quantification of pAMPK/AMPK and pACC/ACC ratios. Phosphorylated protein abundance was normalized to the corresponding total protein abundance. Individual data points represent independent biological samples, and horizontal lines with error bars indicate mean ± standard error of the mean (SEM). Statistical comparisons were performed using ordinary one-way analysis of variance (ANOVA) followed by Šídák’s multiple-comparisons test for three pairwise comparisons (lean versus GNO, lean versus GRO, and GNO versus GRO). Statistical significance is indicated by **P < 0.01, ***P < 0.001, and ****P < 0.0001. GAPDH is shown as a loading-control reference.

### Baseline Hepatic Oxidative Phosphorylation Protein Expression Differs Between GNO and GRO

To characterize baseline hepatic mitochondrial oxidative phosphorylation (OXPHOS) protein expression, we assessed representative subunits of mitochondrial respiratory chain complexes I– V in liver specimens from lean reference controls (n = 5), patients with GLP-1 receptor agonist-naïve obesity (GNO; n = 10), and patients with prior poor response to GLP-1 receptor agonist therapy (GRO; n = 10) (Figure 4A). Quantitative immunoblot analysis demonstrated significant differences in OXPHOS protein expression across the three groups (Figure 4B). Compared with GNO, GRO exhibited significantly lower expression of NDUFS8 (complex I), SDHB (complex II), UQCRC2 (complex III), COX IV (complex IV), and ATP5A (complex V) (all P ≤ 0.0005). NDUFS8, SDHB, and UQCRC2 expression was also significantly lower in GNO than in lean controls (P = 0.0029, P = 0.0106, and P = 0.0009, respectively), whereas ATP5A expression did not differ significantly between these groups (P = 0.0593). COX IV exhibited a distinct expression pattern, with significantly higher abundance in GNO than in lean controls (P = 0.0003) and significantly lower abundance in GRO than in both GNO (P < 0.0001) and lean controls (P = 0.0119). These findings demonstrate differential regulation of hepatic OXPHOS protein abundance in GRO, characterized by reduced expression of representative respiratory-chain subunits relative to GNO across all five complexes.

**Figure 4.**
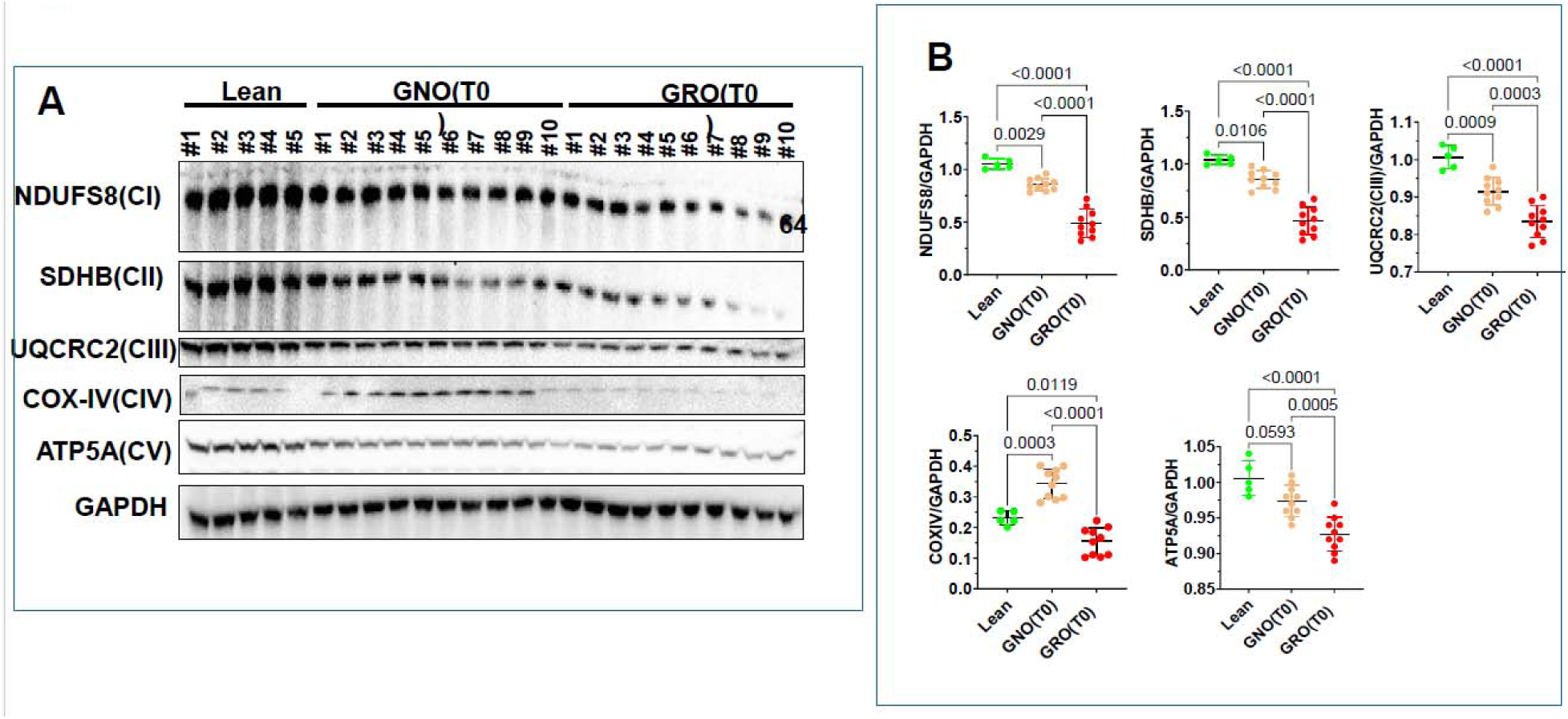
Altered Baseline Hepatic Mitochondrial Oxidative Phosphorylation Protein Expression in Patients With Prior Poor Response to GLP-1 Receptor Agonist Therapy. Baseline hepatic mitochondrial oxidative phosphorylation (OXPHOS) protein expression in lean reference controls (Lean; n = 5), patients with GLP-1 receptor agonist-naïve obesity (GNO; n = 10), and patients with prior poor response to GLP-1 receptor agonist therapy (GRO; n = 10). (A) Representative Western blots showing NDUFS8 (complex I), SDHB (complex II), UQCRC2 (complex III), COX IV (complex IV), and ATP5A (complex V), with GAPDH as the loading control. (B) Quantitative densitometric analyses of NDUFS8, SDHB, UQCRC2, COX IV, and ATP5A protein expressio normalized to GAPDH. Individual data points represent participant-level measurements, and horizontal lines with error bars indicate group mean ± standard error of the mean (SEM). Between-group comparisons were performed using ordinary one-way analysis of variance (ANOVA) followed by Šídák’s multiple-comparisons test for the three pairwise comparisons (Lean versus GNO, Lean versus GRO, and GNO versus GRO). Exact adjusted P values are displayed above the corresponding comparisons.

### Postoperative Weight-Loss Outcomes Following Roux-en-Y Gastric Bypass

Both surgical cohorts demonstrated substantial postoperative weight loss following Roux-en-Y gastric bypass (RYGB), although the magnitude of weight loss was significantly lower among patients with prior poor response to glucagon-like peptide-1 receptor agonist (GLP-1 RA) therapy (GRO) compared with GLP-1 RA-naïve patients (GNO) (Figure 5). At three months, mean percentage total body weight loss (%TWL) was 19.97 ± 1.30% in GRO compared with 23.57 ± 1.10% in GNO, corresponding to a between-group difference of 3.60 percentage points (95% CI, 2.47–4.73; P < 0.0001). At six months, mean %TWL increased to 25.21 ± 2.12% in GRO and 29.51 ± 1.62% in GNO, representing a difference of 4.30 percentage points (95% CI, 2.52–6.08; P < 0.0001). A similar pattern was observed for percentage excess body mass index loss (%EBMIL). At three months, mean %EBMIL was 30.71 ± 1.50% in GRO compared with 34.31 ± 2.13% in GNO, corresponding to a between-group difference of 3.60 percentage points (95% CI, 1.86–5.35; P = 0.0005). By six months, mean %EBMIL increased to 46.50 ± 1.36% in GRO and 50.70 ± 1.36% in GNO, with a between-group difference of 4.20 percentage points (95% CI, 2.92–5.48; P < 0.0001). These findings demonstrate substantial early postoperative weight loss in both cohorts, with significantly greater mean weight-loss magnitudes in GNO than GRO at three and six months (Figure 5; Supplementary Table S1).

**Figure 5.**
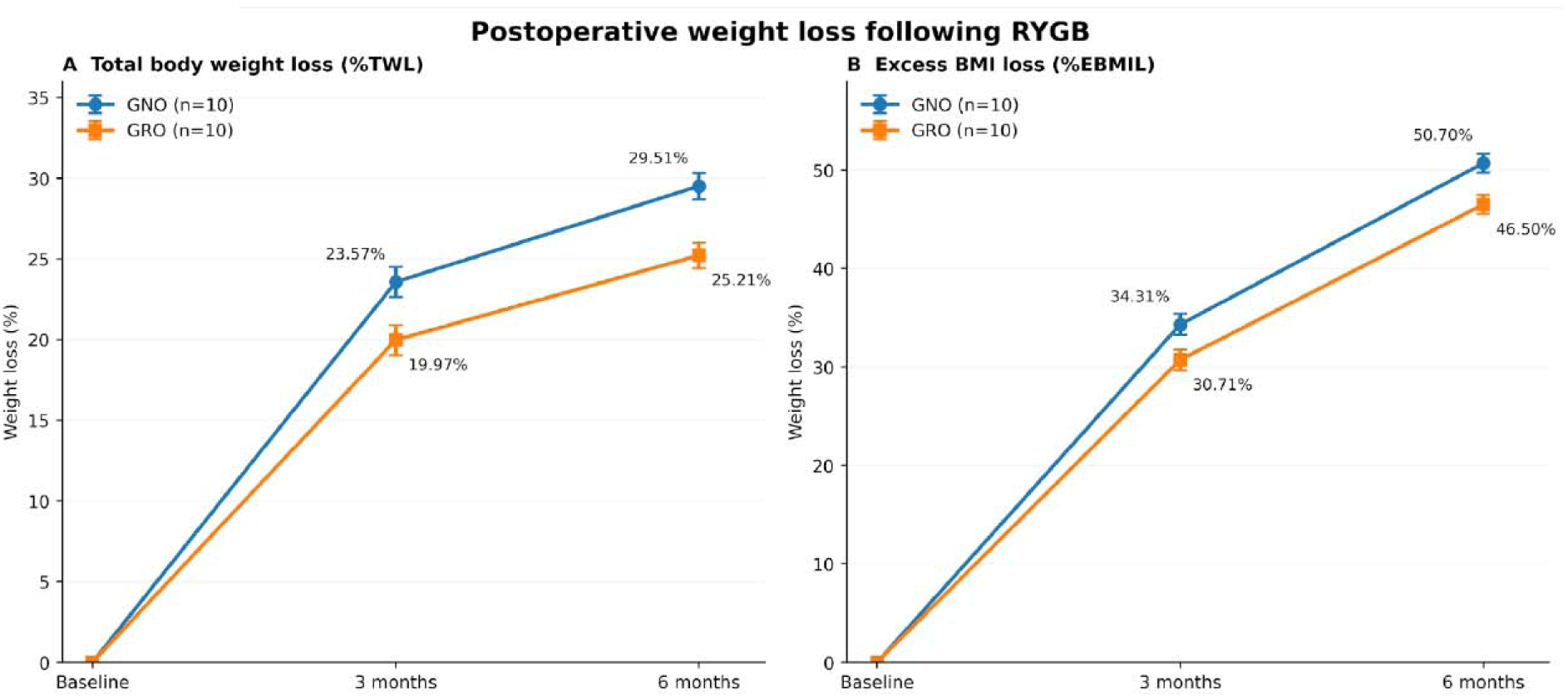
Postoperative Weight-Loss Outcomes Following Roux-en-Y Gastric Bypass in GLP-1 Recepto Agonist–Naïve and Poor-Responder Cohorts. (A) Percentage total body weight loss (%TWL) at baseline, three months, and six months following Roux-en-Y gastric bypass (RYGB) in GLP-1 receptor agonist–naïve patients (GNO; n = 10) and patients with prior poor response to GLP-1 receptor agonist therapy (GRO; n = 10). At three months, mean %TWL was 23.57% in GNO and 19.97% in GRO. By six months, mean %TWL increased to 29.51% and 25.21%, respectively. (B) Percentage excess body mass index loss (%EBMIL) at the correspondin postoperative time points. At three months, mean %EBMIL was 34.31% in GNO and 30.71% in GRO, increasing t 50.70% and 46.50%, respectively, at six months. Data are presented as mean ± standard deviation (SD), with n = 1 participants per cohort. Between-group differences are expressed in percentage points (GNO − GRO), with corresponding 95% confidence intervals (CIs). Statistical comparisons were performed using two-sided Welch’s unpaired t-tests in GraphPad Prism at each postoperative time point, without assuming equal group variances. Analyses were unadjusted for baseline covariates and multiple comparisons.

### Postoperative Hepatic Histological and Mitochondrial Recovery

To evaluate changes in hepatic histological disease activity, intrahepatic lipid accumulation, and mitochondrial ultrastructure following Roux-en-Y gastric bypass (RYGB), we examined paired baseline (T0) and postoperative (T6) liver biopsies from all 20 surgical participants (GNO, n = 10; GRO, n = 10). Postoperative tissue was collected approximately 6–8 months after RYGB during protocolized staged cholecystectomy (Figure 6). Histological examination demonstrated postoperative improvements in hepatic morphology in both cohorts. Representative hematoxylin and eosin (H&E)-stained sections showed reduced hepatocellular vacuolization following RYGB, although residual steatotic changes were more apparent in GRO specimens. Quantitative analysis of changes in the nonalcoholic fatty liver disease activity score (ΔNAS; T6 − T0) demonstrated a greater reduction in GNO compared with GRO (P < 0.0001), indicating a larger improvement in hepatic histological disease activity among GLP-1 receptor agonist-naïve patients. BODIPY 493/503 fluorescence staining similarly demonstrated reductions in intrahepatic neutral lipid accumulation after surgery. The magnitude of reduction in BODIPY-positive area was greater in GNO than GRO (P < 0.0001), consistent with greater postoperative resolution of hepatic lipid accumulation in the GNO cohort. Transmission electron microscopy revealed postoperative changes in mitochondrial ultrastructural organization. GNO specimens demonstrated relatively preserved cristae architecture and mitochondrial matrix organization at T6, whereas GRO specimens continued to exhibit variable cristae fragmentation and structural disorganization. Quantitative analysis using the 0–4 cristae integrity scoring system demonstrated a greater increase in participant-level cristae integrity scores in GNO compared with GRO (P < 0.0001). Across the longitudinal cohort, mitochondrial cristae integrity was assessed using 50 mitochondrial profiles per biopsy, with individual mitochondrial measurements aggregated to obtain participant-level scores. These analyses demonstrated a smaller magnitude of postoperative improvement in GRO than GNO.

**Figure 6.**
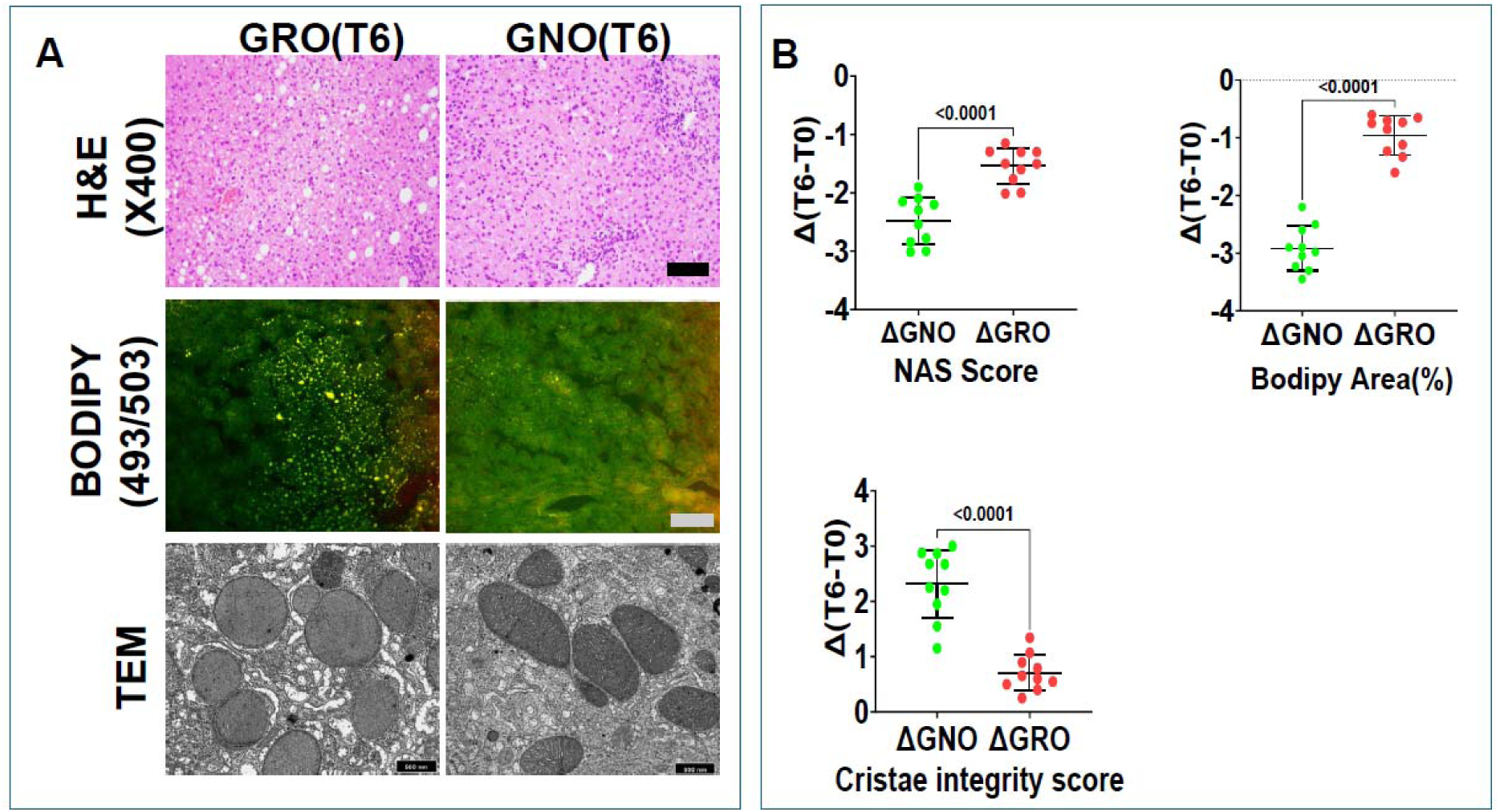
Attenuated Postoperative Hepatic Histological and Mitochondrial Ultrastructural Recovery in Patients With Prior Poor Response to GLP-1 Receptor Agonist Therapy Following Roux-en-Y Gastric Bypass. Representative postoperative hepatic histological, lipid-staining, and mitochondrial ultrastructural images and participant-level changes from baseline (T0) to approximately six months after Roux-en-Y gastric bypass (T6) in GLP-1 receptor agonist-naïve patients (GNO; n = 10) and patients with prior poor response to GLP-1 receptor agonist therapy (GRO; n = 10). (A) Representative hematoxylin and eosin (H&E)-stained liver sections, BODIPY 493/503 fluorescence images, and transmission electron micrographs (TEM) at T6. (B) Participant-level changes in nonalcoholic fatty liver disease activity score (ΔNAS), BODIPY-positive tissue area (ΔBODIPY), an mitochondrial cristae integrity score (Δcristae integrity). Changes were calculated as Δ = T6 − T0. Negative ΔNAS and ΔBODIPY values indicate reductions in disease activity and lipid accumulation, whereas positive Δcristae integrity indicates improved mitochondrial structural preservation. Individual points represent participant-level changes, and horizontal lines with error bars indicate mean ± standard error of the mean (SEM). Between-group comparisons used two-sided unpaired Student’s t-tests assuming equal variances (P < 0.0001 for each outcome). Detailed scoring and image-analysis procedures are provided in the Supplementary Methods.

### Postoperative Hepatic AMPK–ACC Signaling and Longitudinal Changes Following Roux-en-Y Gastric Bypass

To evaluate postoperative changes in hepatic energy-sensing signaling, we assessed AMPK and ACC phosphorylation in liver specimens obtained approximately six months after Roux-en-Y gastric bypass (RYGB) (Figure 7). At T6, the pAMPK/AMPK ratio remained significantly lower in GRO compared with GNO (P < 0.0001). Significant differences were also observed between lean reference controls and both GNO and GRO (P < 0.0001 for each comparison). Similarly, the postoperative pACC/ACC ratio was significantly lower in GRO than GNO (P < 0.0001), with significant differences between lean controls and GNO (P = 0.0145) and between lean controls and GRO (P < 0.0001). Longitudinal analyses demonstrated significantly greater postoperative increases in hepatic pAMPK/AMPK and pACC/ACC ratios in GNO compared with GRO (P = 0.0088 and P < 0.0001, respectively; Figure 7C). Collectively, these findings demonstrate differential postoperative recovery of hepatic AMPK–ACC signaling, with smaller increases in phosphorylation ratios and persistently lower postoperative values in patients with prior poor response to GLP-1 receptor agonist therapy.

**Figure 7.**
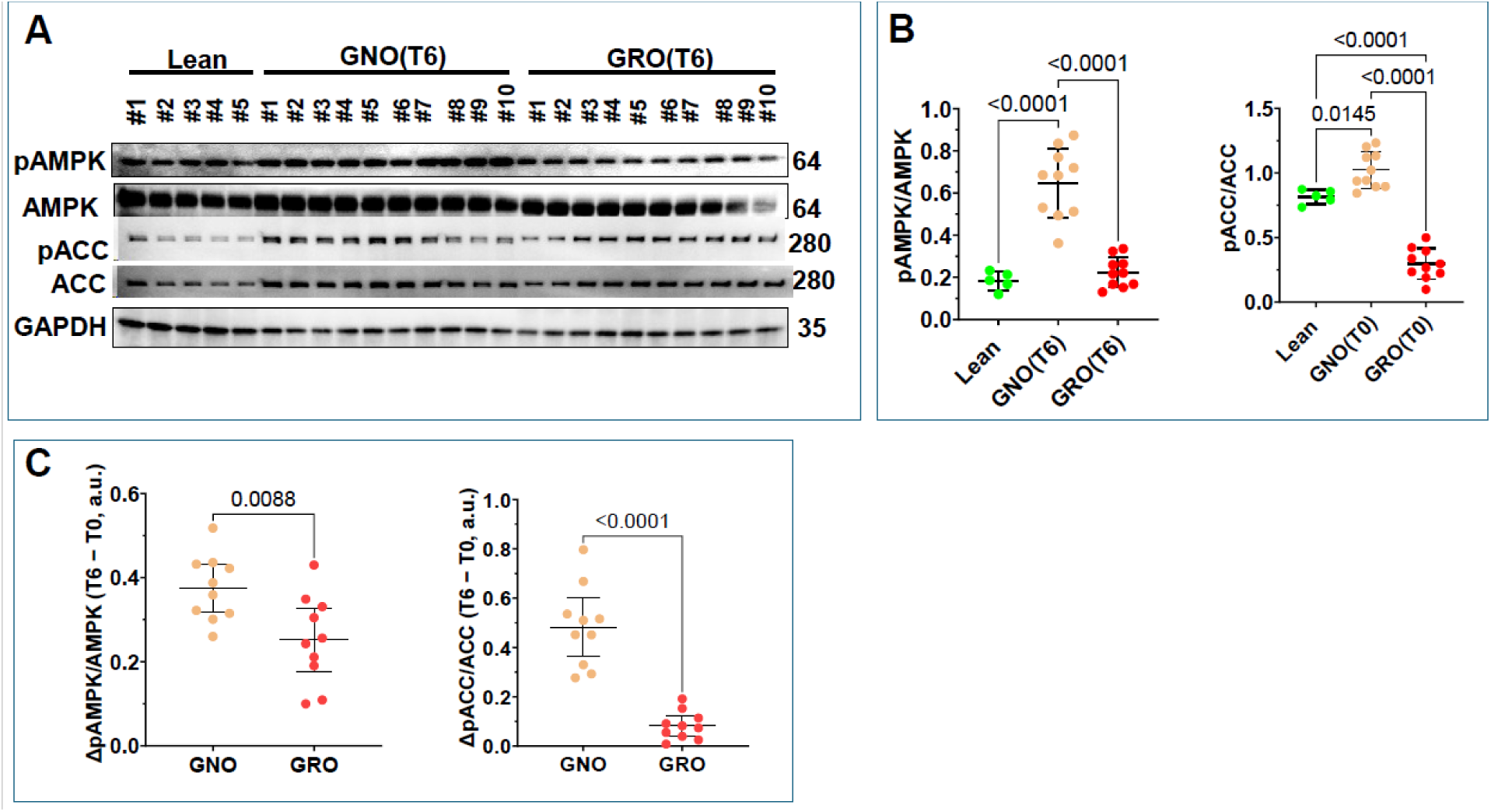
Postoperative Enhancement of Hepatic AMPK–ACC Phosphorylation with Persistent Signalin Differences in Patients With Prior Poor Response to GLP-1 Receptor Agonist Therapy Following Roux-en-Y Gastric Bypass. (A) Representative Western blots showing phosphorylated and total AMP-activated protein kinase (pAMPK and AMPK), phosphorylated and total acetyl-CoA carboxylase (pACC and ACC), and glyceraldehyde-3-phosphate dehydrogenase (GAPDH) in postoperative (T6) liver specimens from lean reference controls (n = 5), GLP-1 receptor agonist-naïve patients (GNO; n = 10), and patients with prior poor response to GLP-1 receptor agonist therapy (GRO; n = 10). (B) Densitometric quantification of pAMPK/AMPK and pACC/ACC ratios at T6. Individual points represent participant-level measurements, with mean ± standard error of the mean (SEM). Cross-sectional comparisons used ordinary one-way analysis of variance (ANOVA) followed by Šídák’s multiple-comparisons test for lean versus GNO, lean versus GRO, and GNO versus GRO; adjusted P values are displayed. (C) Participant-level changes in pAMPK/AMPK and pACC/ACC ratios between baseline (T0) and T6, calculated as Δ = T6 − T0, for GNO and GRO (n = 10 per cohort). Individual points represent participant-level changes with mean ± SEM. Between-group comparisons of Δ values used two-sided Welch’s unpaired t-tests; P values are displayed in panel C. Phosphorylated protein abundance was normalized to corresponding total protein abundance. GAPDH is shown as a loading-control reference. Lean specimens were used for cross-sectional reference comparisons only.

### Postoperative Hepatic Mitochondrial Oxidative Phosphorylation Protein Expression and Longitudinal Changes Following Roux-en-Y Gastric Bypass

To determine whether hepatic mitochondrial oxidative phosphorylation (OXPHOS) protein expression differed between patients with prior poor response to GLP-1 receptor agonist therapy (GRO) and GLP-1 receptor agonist-naïve patients (GNO) following Roux-en-Y gastric bypass (RYGB), we examined representative respiratory-chain proteins in paired baseline (T0) and six-month postoperative (T6) liver specimens (Figure 8). At T6, immunoblot analysis demonstrated differential expression of mitochondrial respiratory-chain proteins between the two surgical cohorts (Figure 8A–B). NDUFS8 (complex I) abundance was comparable between GNO and GRO, with no statistically significant difference in the postoperative cross-sectional comparison. In contrast, SDHB (complex II) expression was significantly lower in GRO compared with GNO (P = 0.0086). Similarly, UQCRC2 (complex III) expression was reduced in GRO relative to GNO (P = 0.0029). ATP5A (complex V) abundance was also lower in GRO compared with GNO (P = 0.0013). The most pronounced difference was observed in cytochrome c oxidase subunit IV (COX IV; complex IV), which exhibited markedly reduced expression in GRO compared with GNO (P < 0.0001). Longitudinal analyses further demonstrated differences in postoperative changes in OXPHOS protein abundance between the two cohorts (Figure 8C). Changes in NDUFS8/GAPDH were greater in GRO than GNO (P = 0.0038), despite comparable absolute protein abundance at T6. In contrast, the postoperative increase in SDHB/GAPDH was significantly greater in GNO compared with GRO (P < 0.0001). Similarly, GNO demonstrated greater postoperative changes in UQCRC2/GAPDH (P < 0.0001), COX IV/GAPDH (P < 0.0001), and ATP5A/GAPDH (P < 0.0001) compared with GRO. Together, these findings demonstrate heterogeneous postoperative remodeling of hepatic mitochondrial OXPHOS protein expression following RYGB. Although complex I abundance was comparable between cohorts at T6 and exhibited a greater longitudinal increase in GRO, the remaining respiratory-chain proteins demonstrated lower postoperative abundance and smaller changes in GRO relative to GNO. This pattern suggests that hepatic mitochondrial protein-expression responses following RYGB differ according to prior GLP-1 receptor agonist treatment-response phenotype. However, differences in respiratory-chain protein abundance do not directly establish alterations in mitochondrial respiratory activity or ATP-generating capacity.

**Figure 8.**
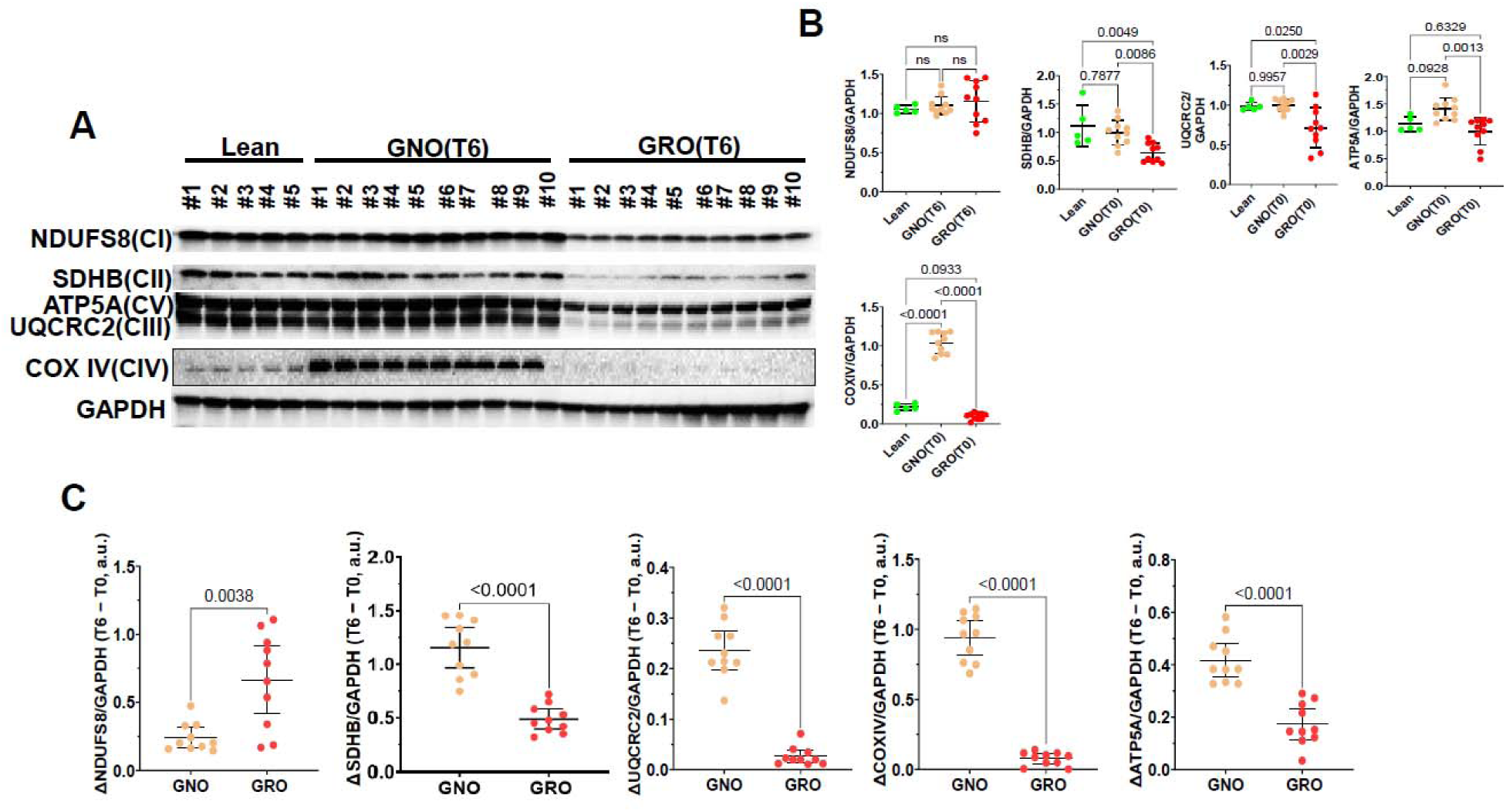
Differential Postoperative Hepatic Mitochondrial Oxidative Phosphorylation Protein Expression and Longitudinal Remodeling in Patients with Prior Poor Response to GLP-1 Receptor Agonist Therap Following Roux-en-Y Gastric Bypass. (A) Representative Western blots showing mitochondrial oxidative phosphorylation (OXPHOS) proteins in liver specimens obtained approximately six months after Roux-en-Y gastric bypass (T6). Samples include lean physiological reference controls (n = 5), GLP-1 receptor agonist-naïve patients (GNO; n = 10), and patients with prior poor response to GLP-1 receptor agonist therapy (GRO; n = 10). Th analyzed proteins include NDUFS8 (complex I), SDHB (complex II), UQCRC2 (complex III), COX IV (complex IV), and ATP5A (complex V). GAPDH was used as a loading-control reference. (B) Densitometric quantification of individual OXPHOS proteins normalized to GAPDH at T6. Individual data points represent independent biological samples, and group summaries indicate mean ± standard error of the mean (SEM). Statistical comparisons were performed using ordinary one-way analysis of variance (ANOVA) followed by Šídák’s multiple-comparisons testfor lean versus GNO, lean versus GRO, and GNO versus GRO. (C) Longitudinal changes in GAPDH-normalize mitochondrial OXPHOS protein abundance between baseline (T0) and postoperative follow-up (T6). Changes were calculated for each participant as Δ = T6 − T0. Individual data points represent participant-level changes (n = 10 per surgical cohort), and group summaries indicate mean ± SEM. Between-group comparisons of longitudinal changes were performed using two-sided Welch’s unpaired t-tests without assuming equal group variances. Lean reference controls were included in cross-sectional comparisons only and were not included in longitudinal analyses.

## Discussion

In this prospective observational study, patients with prior poor response to glucagon-like peptide-1 receptor agonist (GLP-1 RA) therapy exhibited a distinct hepatic metabolic phenotype compared with GLP-1 RA-naïve patients undergoing Roux-en-Y gastric bypass (RYGB). Despite broadly comparable baseline anthropometric and metabolic characteristics, the GRO cohort demonstrated greater hepatic lipid accumulation, more pronounced mitochondrial ultrastructural abnormalities, and lower AMPK–ACC phosphorylation ratios. Both cohorts subsequently achieved substantial postoperative weight loss and improvements in hepatic histological and mitochondrial parameters; however, the magnitude of tissue-level recovery was generally smaller in GRO. Persistent differences in postoperative AMPK–ACC phosphorylation and complex-specific oxidative phosphorylation (OXPHOS) protein remodeling further suggest that hepatic metabolic characteristics remain distinguishable between these clinical phenotypes after surgery. These findings identify an association between prior pharmacologic response history and hepatic metabolic heterogeneity, without establishing a causal relationship.

The clinical relevance of these findings is increasing as GLP-1-based therapies become established components of contemporary obesity management. Obesity affects more than one billion individuals worldwide, and the expanding therapeutic role of incretin-based medications has substantially changed the treatment landscape. Although semaglutide, liraglutide, and newer incretin-based therapies have demonstrated meaningful efficacy, individual weight-loss responses remain heterogeneous, and a proportion of patients do not achieve clinically meaningful weight reduction despite adequate treatment exposure.^3, 4^ As pharmacotherapy becomes more widely incorporated into longitudinal obesity care, metabolic and bariatric surgery will increasingly be considered in patients with prior GLP-1-based treatment exposure, including those with inadequate pharmacologic response. However, the biological characteristics of patients with inadequate GLP-1 receptor agonist response remain poorly defined. In particular, whether poor pharmacologic weight-loss response is associated with distinct hepatic metabolic abnormalities and whether these abnormalities persist following metabolic surgery remain important unanswered questions. Existing investigations have largely emphasized clinical weight-loss efficacy and metabolic outcomes, with limited longitudinal human tissue evidence addressing these relationships.^16^ A distinguishing strength of the present study is the prospective collection of paired liver biopsies at the time of RYGB and approximately six months postoperatively, enabling integrated assessment of hepatic histology, mitochondrial ultrastructure, and molecular signaling in patients stratified by prior pharmacologic response. This approach provides tissue-level insights beyond conventional clinical outcomes and addresses an emerging knowledge gap as incretin-based therapies become increasingly integrated into obesity management. Nevertheless, the observed associations do not establish a mechanism of GLP-1 receptor resistance or distinguish poor-response-associated biology from the effects of prior treatment exposure.

The greater baseline hepatic histological disease activity and mitochondrial structural abnormalities observed in GRO are noteworthy because the cohorts had similar measured BMI, glycemic indices, and insulin resistance. These findings suggest that conventional clinical parameters may not fully capture hepatic metabolic heterogeneity in severe obesity. Mitochondrial cristae architecture is closely linked to the organization of the respiratory machinery and maintenance of cellular energy metabolism.^17^ The combination of greater lipid accumulation, disrupted cristae, and altered respiratory-chain protein abundance in GRO is therefore consistent with differences in mitochondrial homeostasis. However, mitochondrial morphology and protein abundance cannot independently establish impaired oxidative phosphorylation or respiratory capacity.

Altered AMPK–ACC phosphorylation provides a plausible molecular context for these observations. AMPK is a central regulator of cellular energy homeostasis that coordinates anabolic and catabolic pathways in response to energetic stress.^9, 10^ One of its principal downstream targets, acetyl-CoA carboxylase (ACC), regulates malonyl-CoA production and thereby influences hepatic lipid synthesis and mitochondrial fatty acid oxidation.^18^ AMPK-mediated phosphorylation inhibits ACC activity, providing a mechanistic link between energy sensing and lipid metabolism. In the present study, reduced baseline pAMPK/AMPK and pACC/ACC ratios in GRO occurred alongside greater hepatic lipid accumulation and mitochondrial structural abnormalities. This concordance suggests altered AMPK-associated metabolic signaling in patients with prior poor pharmacologic response. Nevertheless, phosphorylation ratios do not directly measure enzymatic activity, malonyl-CoA concentrations, or fatty acid oxidation, and these relationships remain mechanistic hypotheses rather than established causal pathways.

Both cohorts experienced substantial postoperative weight loss, with mean six-month total body weight loss of 29.51% in GNO and 25.21% in GRO. These findings are clinically relevant because inadequate response to GLP-1 RA therapy did not preclude substantial benefit from RYGB. Unlike pharmacologic GLP-1 receptor stimulation, RYGB produces coordinated physiological changes involving nutrient delivery, gastrointestinal hormone secretion, neural signaling, and systemic metabolic regulation.^19^ Thus, the observed surgical response in GRO is biologically plausible and reinforces the distinction between inadequate pharmacologic weight-loss response and resistance to metabolic surgery. The numerical difference in postoperative weight loss between cohorts should nevertheless be interpreted cautiously because the study was not designed to establish prior GLP-1 RA response as an independent predictor of surgical outcomes.

The paired liver biopsy design enabled direct assessment of hepatic remodeling following RYGB. Both cohorts demonstrated reductions in histological disease activity and intrahepatic lipid accumulation, accompanied by improvements in mitochondrial cristae integrity, consistent with previous longitudinal studies reporting histological improvement following bariatric surgery.^14, 15^ However, the magnitude of hepatic histological and ultrastructural recovery was consistently smaller in GRO despite substantial postoperative weight loss in both cohorts. This finding suggests that weight reduction and tissue-level metabolic remodeling may represent related but distinct dimensions of response to metabolic surgery. Importantly, the observed differences in hepatic recovery may reflect baseline disease severity, differences in postoperative weight loss, prior pharmacologic exposure, or other unmeasured factors rather than a direct consequence of GLP-1 RA nonresponse. Whether these differences persist independently of weight-loss magnitude or influence longer-term hepatic outcomes remains to be determined.

Longitudinal molecular analyses further demonstrated that postoperative hepatic remodeling was not uniform across pathways or respiratory-chain complexes. Although AMPK and ACC phosphorylation ratios increased in both surgical cohorts, postoperative ratios remained lower in GRO. Similarly, mitochondrial OXPHOS proteins exhibited heterogeneous responses: NDUFS8 showed a greater longitudinal increase in GRO and comparable abundance between cohorts at six months, whereas SDHB, UQCRC2, COX IV, and ATP5A remained lower in GRO and generally demonstrated smaller postoperative changes. These findings suggest that mitochondrial protein remodeling after RYGB is complex-specific rather than a generalized restoration of respiratory-chain abundance. The observed patterns may reflect differences in mitochondrial content, protein turnover, or respiratory-chain regulation, but direct functional measurements would be required to distinguish these possibilities.

The clinical implications of these findings are particularly relevant given the expanding use of incretin-based pharmacotherapy and the increasing likelihood that patients with prior inadequate pharmacologic response will be evaluated for metabolic surgery.^16, 20^ Although GLP-1 receptor agonists have demonstrated beneficial effects on hepatic steatohepatitis in selected populations, the biological characteristics underlying variability in weight-loss response remain incompletely understood.^13, 21^ Importantly, GRO was defined by inadequate weight loss despite sustained therapeutic exposure, rather than demonstrated hepatic resistance to GLP-1 signaling. The observed hepatic abnormalities may therefore reflect pre-existing metabolic heterogeneity, prior pharmacologic exposure, or other unmeasured factors. Nevertheless, our findings suggest that pharmacologic response history may provide clinically relevant context beyond BMI and conventional metabolic indices when investigating postoperative hepatic recovery. Whether these characteristics can ultimately inform individualized treatment selection, postoperative surveillance, or long-term metabolic outcomes require validation in larger cohorts incorporating GLP-1 RA responders and direct assessments of mitochondrial function.

The study has several strengths, including its prospective design, defined pharmacologic exposure criteria, standardized surgical procedure, and availability of paired human liver biopsies from all 20 surgical participants. Integration of histological scoring, fluorescence-based lipid quantification, mitochondrial ultrastructure, and protein-expression analyses provided complementary assessments of hepatic remodeling. However, the small, single-center cohort limits statistical precision and adjustment for potential confounding. The absence of a GLP-1 RA responder comparator prevents separation of poor-response-associated characteristics from prior treatment exposure. The lean reference cohort was small and clinically distinct, and the six-month follow-up does not establish long-term durability. Furthermore, OXPHOS protein abundance and AMPK–ACC phosphorylation are indirect molecular measurements rather than functional assessments of mitochondrial respiration or metabolic flux. Multiple exploratory endpoints also increase the possibility of chance findings. These limitations preclude causal or predictive conclusions.

In conclusion, prior poor response to GLP-1 RA therapy was associated with greater baseline hepatic histological abnormalities, altered AMPK–ACC phosphorylation, and differences in mitochondrial ultrastructure and respiratory-chain protein abundance among patients undergoing RYGB. Although both surgical cohorts achieved substantial postoperative weight loss and hepatic improvement, differences in tissue-level recovery and molecular profiles remained evident at six months. These findings support further investigation of hepatic energy sensing and mitochondrial homeostasis as potential contributors to biological heterogeneity in obesity treatment response.

## Data Availability

All data produced in the present work are contained in the manuscript

## List of Abbreviations

ACC: Acetyl-CoA carboxylase
AMPK: AMP-activated protein kinase
ANOVA: Analysis of variance
ATP5A: ATP synthase F1 subunit alpha
BMI: Body mass index
BODIPY: Boron-dipyrromethene
CI: Confidence interval
COMIRB: Colorado Multiple Institutional Review Board
COX IV: Cytochrome c oxidase subunit IV
GAPDH: Glyceraldehyde-3-phosphate dehydrogenase
GLP-1: Glucagon-like peptide-1
GLP-1 RA: Glucagon-like peptide-1 receptor agonist
GNO: GLP-1 receptor agonist-naïve obesity
GRO: GLP-1 receptor agonist–refractory obesity
H&E: Hematoxylin and eosin
HbA1c: Glycated hemoglobin
HOMA-IR: Homeostatic model assessment of insulin resistance
IQR: Interquartile range
MASLD: Metabolic dysfunction-associated steatotic liver disease
NAS: Nonalcoholic fatty liver disease activity score
NDUFS8: NADH:ubiquinone oxidoreductase core subunit S8
OXPHOS: Oxidative phosphorylation
pACC: Phosphorylated acetyl-CoA carboxylase
pAMPK: Phosphorylated AMP-activated protein kinase
RYGB: Roux-en-Y gastric bypass
SD: Standard deviation
SDHB: Succinate dehydrogenase subunit B
SEM: Standard error of the mean
STROBE: Strengthening the Reporting of Observational Studies in Epidemiology
T0: Baseline liver biopsy time point
T6: Postoperative liver biopsy time point (approximately six months)
TEM: Transmission electron microscopy
%EBMIL: Percentage excess body mass index loss
%TWL: Percentage total body weight loss
UQCRC2: Ubiquinol-cytochrome c reductase core protein 2

## References

1. Liu Z, Yu S, Jin X, et al. The Clinical Application of GLP-1RAs and GLP-1/GIP Dual Receptor Agonists Based on Pharmacological Mechanisms: A Review. Drug Des Devel Ther. 2025;19:10383–10409.

2. Scheen AJ, Paquot N. [Potentiation of weight reduction with GLP-1 receptor agonists]. Rev Med Suisse. 2021;17(747):1405–1410.

3. Wilding JPH, Batterham RL, Calanna S, et al. Once-Weekly Semaglutide in Adults with Overweight or Obesity. N Engl J Med. 2021;384(11):989–1002.

4. Jastreboff AM, Aronne LJ, Ahmad NN, et al. Tirzepatide once weekly for the treatment of obesity. New England Journal of Medicine. 2022;387(3):205–216.

5. Klein MP, Akturk HK, Snell-Bergeon JK, Shah VN. Reduced Efficacy of Glucagon-Like Peptide-1 Receptor Agonists Therapy in People With Type 1 Diabetes and Genetic Forms of Obesity. J Diabetes Sci Technol. 2025;19(2):297–303.

6. Lutz TA, Bueter M. Physiological Mechanisms behind Roux-en-Y Gastric Bypass Surgery. Digestive Surgery. 2014;31(1):13–24.

7. Abdeen G, le Roux CW. Mechanism Underlying the Weight Loss and Complications of Roux-en-Y Gastric Bypass. Review. Obes Surg. 2016;26(2):410-421.

8. Klein S, Mittendorfer B, Eagon JC, et al. Gastric Bypass Surgery Improves Metabolic and Hepatic Abnormalities Associated With Nonalcoholic Fatty Liver Disease. Gastroenterology. 2006;130(6):1564–1572.

9. Hardie DG, Ross FA, Hawley SA. AMPK: a nutrient and energy sensor that maintains energy homeostasis. Nature Reviews Molecular Cell Biology. 2012;13(4):251–262.

10. Herzig S, Shaw RJ. AMPK: guardian of metabolism and mitochondrial homeostasis. Nature Reviews Molecular Cell Biology. 2018;19(2):121–135.

11. Viollet B, Foretz M, Guigas B, et al. Activation of AMP-activated protein kinase in the liver: a new strategy for the management of metabolic hepatic disorders. J Physiol. 2006;574(Pt 1):41–53.

12. Koliaki C, Szendroedi J, Kaul K, et al. Adaptation of Hepatic Mitochondrial Function in Humans with Non-Alcoholic Fatty Liver Is Lost in Steatohepatitis. Cell Metabolism. 2015;21(5):739–746.

13. Newsome PN, Buchholtz K, Cusi K, et al. A Placebo-Controlled Trial of Subcutaneous Semaglutide in Nonalcoholic Steatohepatitis. N Engl J Med. 2021;384(12):1113–1124.

14. Lassailly G, Caiazzo R, Buob D, et al. Bariatric Surgery Reduces Features of Nonalcoholic Steatohepatitis in Morbidly Obese Patients. Gastroenterology. 2015;149(2):379–388; quiz e315-376.

15. Lassailly G, Caiazzo R, Ntandja-Wandji LC, et al. Bariatric Surgery Provides Long-term Resolution of Nonalcoholic Steatohepatitis and Regression of Fibrosis. Gastroenterology. 2020;159(4):1290–1301.e1295.

16. AbuHasan Q, Funk LM, Holl JL, et al. Preoperative Glucagon-Like Peptide-1 Receptor Agonist Utilization and Association with Bariatric Surgery Outcomes. Obesity Surgery. 2025;35(2):556–560.

17. Friedman JR, Nunnari J. Mitochondrial form and function. Nature. 2014;505(7483):335–343.

18. Fullerton MD, Galic S, Marcinko K, et al. Single phosphorylation sites in Acc1 and Acc2 regulate lipid homeostasis and the insulin-sensitizing effects of metformin. Nature Medicine. 2013;19(12):1649–1654.

19. Sinclair P, Brennan DJ, le Roux CW. Gut adaptation after metabolic surgery and its influences on the brain, liver and cancer. Nature Reviews Gastroenterology & Hepatology. 2018;15(10):606–624.

20. Eisenberg D, Shikora SA, Aarts E, et al. 2022 American Society of Metabolic and Bariatric Surgery (ASMBS) and International Federation for the Surgery of Obesity and Metabolic Disorders (IFSO) Indications for Metabolic and Bariatric Surgery. Obesity Surgery. 2023;33(1):3-14.

21. Armstrong MJ, Gaunt P, Aithal GP, et al. Liraglutide safety and efficacy in patients with non-alcoholic steatohepatitis (LEAN): a multicentre, double-blind, randomised, placebo-controlled phase 2 study. The Lancet. 2016;387(10019):679–690.

