## Supplementary Methods for "Hepatic Mitochondrial Alterations in GLP-1 Poor Responders Undergoing Roux-en-Y Gastric Bypass"

**S1. Study Eligibility and Pharmacologic Exposure Assessment**

Participants were screened during preoperative evaluation for primary Roux-en-Y gastric bypass (RYGB). Eligible participants were adults aged at least 18 years who met the study's BMI and comorbidity criteria.

Patients classified as having poor response to GLP-1 receptor agonist therapy (GRO) were required to have less than 5% total body weight loss following an adequate treatment course consisting of standard dose escalation and at least six consecutive months at the prescribed maintenance dose.

Medication histories were reviewed to document the GLP-1 receptor agonist used, maintenance dose, total treatment duration, duration at maintenance dosing, treatment adherence, and weight-loss response. Semaglutide and liraglutide exposure was recorded separately. Participants who were unable to achieve or maintain the required treatment dose because of intolerance were excluded from the GRO cohort.

Medication exposure and response were assessed using electronic health records, prescription histories, and clinician documentation. Total treatment duration and duration at the maintenance dose were recorded separately.

**S2. Surgical Procedures and Tissue Collection**

All surgical participants underwent primary RYGB with a standardized operative configuration consisting of a 120-cm biliopancreatic limb and a 100-cm Roux limb.

Staged cholecystectomy was planned approximately 6–8 months after RYGB according to the institutional surgical pathway. Patients were counseled regarding gallstone-related complications after substantial postoperative weight loss and the potential challenges of biliary interventions following gastrointestinal reconstruction. The planned procedure and longitudinal research tissue collection were incorporated into the institutional informed-consent process.

Baseline liver biopsies (T0) were collected during the index RYGB. Postoperative biopsies (T6) were obtained during the subsequent staged cholecystectomy.

Twenty surgical participants contributed paired biopsies, yielding 40 surgical biopsy specimens. Specimens were allocated for histopathology, lipid fluorescence staining, TEM, and molecular analyses.

Five lean reference participants contributed liver specimens for physiological comparisons. Lean specimens were not included in paired postoperative analyses.

**S3. Histopathological Assessment and NAS Scoring**

Liver tissue sections were stained with hematoxylin and eosin and evaluated using the NASH Clinical Research Network histological scoring system described by Kleiner et al.

The NAFLD Activity Score (NAS) was calculated from three components:

| **Component** | **Score** | **Criteria** |
| --- | --- | --- |
| Steatosis | 0–3 | 0: <5%; 1: 5–33%; 2: >33–66%; 3: >66% of hepatocytes |
| Lobular inflammation | 0–3 | 0: none; 1: <2 foci; 2: 2–4 foci; 3: >4 foci per 200× field |
| Hepatocellular ballooning | 0–2 | 0: none; 1: few ballooned cells; 2: many or prominent ballooned cells |

The component scores were summed to obtain a composite NAS ranging from 0 to 8.

Fibrosis was assessed separately and was not included in the NAS.

Histological assessment was performed by a hepatopathologist blinded to clinical group assignment. Baseline and postoperative specimens were evaluated using identical scoring criteria.

**S4. BODIPY 493/503 Fluorescence Imaging and Quantification**

Intrahepatic neutral lipid accumulation was assessed using BODIPY 493/503 fluorescence staining.

For each biopsy, 10 spatially distinct, nonoverlapping fluorescence microscopy fields were evaluated. Digital images were analyzed using Fiji/ImageJ.

Tissue regions of interest were identified to distinguish evaluable tissue from background and non-tissue regions. BODIPY-positive fluorescence was quantified as the percentage of total evaluable tissue area exhibiting positive signal.

BODIPY-positive area (%) = (BODIPY-positive tissue area / Total evaluable tissue area) × 100.

Measurements from the 10 fields were averaged to generate one participant-level value per biopsy.

Longitudinal changes were calculated as the postoperative percentage-positive area minus the corresponding baseline percentage-positive area. Changes were expressed in percentage points.

**S5. Transmission Electron Microscopy and Cristae Integrity Scoring**

Liver specimens were fixed in glutaraldehyde and processed for transmission electron microscopy (TEM).

For each biopsy, 10 spatially distinct, nonoverlapping TEM fields were evaluated. Five mitochondrial profiles were scored per field, yielding 50 mitochondrial observations per biopsy.

Across the 40 surgical biopsy specimens, the sampling protocol comprised 400 TEM fields and 2,000 mitochondrial profiles.

Mitochondrial cristae integrity was evaluated using a semiquantitative scoring system ranging from 0 to 4:

| **Score** | **Morphological criteria** |
| --- | --- |
| 0 | Severe cristae disruption or near-complete loss |
| 1 | Marked fragmentation, disorganization, or substantial cristae loss |
| 2 | Moderate abnormalities with partial cristae preservation |
| 3 | Predominantly preserved cristae with mild structural irregularities |
| 4 | Intact, regularly organized cristae with preserved mitochondrial architecture |

Individual mitochondrial scores were averaged across the 50 profiles to generate one participant-level cristae integrity score per biopsy.

The postoperative change in cristae integrity was calculated as the T6 score minus the corresponding T0 score. Positive values indicated improvement in mitochondrial structural preservation.

**S6. Western Blot Analysis and Densitometry**

Frozen liver tissue was homogenized in radioimmunoprecipitation assay (RIPA) buffer containing protease and phosphatase inhibitors.

Equal quantities of total protein were separated by sodium dodecyl sulfate–polyacrylamide gel electrophoresis (SDS-PAGE) and transferred to polyvinylidene difluoride membranes.

Immunoblotting was performed for phosphorylated and total AMPK and ACC, together with representative mitochondrial oxidative phosphorylation proteins.

The mitochondrial targets included NDUFS8 (complex I), SDHB (complex II), UQCRC2 (complex III), COX IV (complex IV), and ATP5A (complex V).

GAPDH was used as a loading-control reference.

Protein bands were quantified by densitometry. Phosphorylated protein measurements were normalized to their corresponding total protein measurements. Mitochondrial OXPHOS proteins were normalized to GAPDH.

Where samples were distributed across multiple membranes, inter-blot reference samples were used to support normalization.

Five independent lean specimens and 10 specimens from each surgical cohort were included in baseline Western blot analyses.

**S7. Clinical Outcome Definitions**

Percentage total body weight loss (%TWL) was calculated as:

%TWL = [(Preoperative weight − Postoperative weight) / Preoperative weight] × 100.

The preoperative reference weight was the weight recorded at the time of RYGB.

Percentage excess BMI loss (%EBMIL) was evaluated as a secondary surgical outcome.

Clinical outcomes were assessed at three and six months after surgery.

**S8. Statistical Analysis**

Baseline clinical variables were summarized using descriptive statistics. Continuous variables were compared using Welch's independent-samples t-test, and categorical variables were compared using Fisher's exact test.

Baseline histological and ultrastructural outcomes were analyzed using ordinary one-way ANOVA followed by Šídák's multiple-comparisons test for the three pairwise comparisons (lean versus GNO, lean versus GRO, and GNO versus GRO). Baseline AMPK–ACC phosphorylation ratios and OXPHOS protein abundance were analyzed using ordinary one-way ANOVA followed by Šídák's multiple-comparisons test for the three pairwise comparisons (lean versus GNO, lean versus GRO, and GNO versus GRO).

For longitudinal hepatic outcomes, postoperative changes were calculated separately for each participant as Δ = T6 − T0.

Between-group differences in participant-level changes in NAS, BODIPY-positive area, and cristae integrity (GNO versus GRO) were evaluated using two-sided unpaired Student's t-tests assuming equal variances. Between-group differences in participant-level changes in AMPK–ACC phosphorylation ratios and OXPHOS protein abundance were evaluated using two-sided Welch's unpaired t-tests without assuming equal variances. Postoperative percentage total weight loss and percentage excess BMI loss were compared between groups at three and six months using two-sided Welch's unpaired t-tests.

Within-participant measurements from multiple microscopic fields or mitochondrial profiles were summarized before statistical testing to avoid pseudoreplication.

Exploratory associations between baseline hepatic protein expression and six-month %TWL were evaluated using Spearman rank correlations. Pooled analyses and within-group analyses were distinguished where applicable.

All tests were two-sided, with nominal statistical significance defined as P < 0.05. The exploratory nature of the molecular analyses and the potential influence of multiple comparisons were considered when interpreting the findings.
